# Dual-outcome Prediction of Post-Ischemic Stroke Epilepsy and Mortality Using Multimodal Quantitative Biomarkers

**DOI:** 10.1101/2025.09.22.25335736

**Authors:** Yilun Chen, Alexandria L. Soto, Tejaswi D. Sudhakar, Adeel Zubair, Haoqi Sun, Jin Jing, Wendong Ge, Lucas Loman, Adithya Sivaraju, Nils Petersen, Lawrence J. Hirsch, Hal Blumenfeld, Sahar F. Zafar, Aaron F. Struck, Kevin N. Sheth, Emily J. Gilmore, M. Brandon Westover, Jennifer A. Kim

**Affiliations:** Department of Neurology, Yale University School of Medicine, 333 Cedar Street, New Haven, CT 06510, USA; Department of Neurology, Massachusetts General Hospital, 55 Fruit St, Boston, MA 02114, USA; Department of Neurology, University of Wisconsin School of Medicine and Public Health, 750 Highland Ave, Madison, WI 53726, USA; Yale Center for Brain & Mind Health, Yale University School of Medicine, 333 Cedar Street, New Haven, CT 06510, USA

## Abstract

**Background and Objectives:** Post-ischemic stroke epilepsy (PISE) reduces quality of life, and early risk prediction can guide prevention strategies and anti-epileptogenesis treatment trials. Stroke severity predicts both PISE and mortality, and ignoring mortality can overestimate epilepsy risk. We sought to enhance PISE risk stratification by modeling death as a competing outcome, integrating quantitative clinical, neuroimaging, and electroencephalography (EEG) biomarkers to distinguish shared and distinct predictors of epilepsy and mortality.

**Methods:** We developed a PISE prediction model using retrospective data from Yale-New Haven Hospital. The training cohort included patients from 2014–2020; the testing cohort from 2021–2022. Eligible patients were adults with acute ischemic stroke who underwent neuroimaging and EEG monitoring <7 days post-stroke and had follow-up >7 days.

**Results:** Of 280 patients, 53 developed PISE first, 104 died first, and the rest were censored. Quantitative PISE biomarkers included greater 72h stroke severity (HR_Δ3_ [95%CI], 1.2 [1.1-1.4]), infarct volume (HR_Δ10mL_, 1.06 [1.04-1.08]), EEG epileptiform abnormality burden (HR_Δ10%_, 1.2 [1.1-1.3]), and EEG power asymmetries (HR_Δ10%_, 2.0 [1.4-2.9]). Death predictors included older age (HR_Δ10years_, 1.7 [1.4-2.0]), worse pre-stroke functional status (HR, 1.4 [1.2-1.7]), atrial fibrillation history (HR, 2.4 [1.6-3.7]), cardioembolism etiology (HR, 1.9 [1.2-3.0]), anterior cerebral artery involvement (HR, 2.2 [1.2-3.7]), and greater EEG global theta-band powers (HR_Δ10µV_, 6.2 [2.3-17]). Our model, CRIME_PISE_, integrating these features, allows prediction of PISE-first and death-first risk scores with AUC of 0.72 (95%CI, 0.60-0.83) and 0.79 (0.72-0.85), respectively. Compared with the benchmark SeLECT model, CRIME_PISE_ better predicted PISE in patients with ≥4 SeLECT points (AUC, 0.72 vs 0.58) but not those with <4 points (AUC, 0.33 vs 0.52). In the testing cohort, CRIME_PISE_ identified a more selective group (n=18 vs 44 per SeLECT) with a higher PISE rate (39% vs 20%) and a lower mortality rate (22% vs 45%).

**Discussion:** CRIME_PISE_ enhances PISE prediction by accounting for mortality as a competing outcome and incorporating multimodal quantitative biomarkers. Because its benefits over SeLECT are most pronounced in high-risk patients, a two-stage approach—SeLECT screening followed by CRIME_PISE_ in SeLECT-positive cases—may better target candidates for anti-epileptogenesis trials by prioritizing patients likely to survive long-term and develop epilepsy.

## Introduction

Acute ischemic stroke (AIS) causes over a million deaths annually,^1^ and survivors often face complications like seizures. Acute post-AIS seizures are associated with higher in-hospital costs and increased mortality.^2,3^ Long-term unprovoked seizures, called post-ischemic stroke epilepsy (PISE),^4^ reduce patients’ quality of life and are difficult to treat.^5^ The development of PISE treatment is hindered by high trial costs due to the low incidence of PISE. An accurate PISE risk stratification model can reduce these costs by targeting trial enrollments to high-risk patients.^6^

To be clinically useful, a PISE risk model must identify a subgroup of patients with sufficiently high positive predictive value (PPV) to justify intervention. This subgroup should also be large enough to enable meaningful clinical application and trial enrollment. Yet, the benchmark PISE prediction model, SeLECT (Severity of the stroke, Large artery atherosclerosis, Early seizure, Cortical involvement, Territory of medial cerebral artery)^7–9^ has a PPV of only 11% at one-year post-stroke.^4,7,10^ The majority of the remaining 89% are likely false positives, either because *seizures never occur or because patients die before developing epilepsy*.

Conceptually, two complementary strategies can be adopted to boost the PPV of PISE prediction models.

One strategy is to incorporate additional PISE biomarkers.^5,7^ Prior studies suggest that in-hospital stroke severity score,^11^ stroke size,^12–15^ and the presence of electroencephalography (EEG) abnormalities^16–21^ were predictive of PISE. However, most prior approaches relied on binary or qualitative measures. Algorithmically derived quantitative metrics, especially for continuous EEG recordings, improved scalability and provided additive benefits for post-injury epilepsy prediction as compared to expert-reviewed qualitative features.^22,23^ Yet, these quantitative biomarkers have not yet been integrated into a unified PISE prediction model.

Most existing PISE prediction models include PISE predictors such as stroke severity, which also correlate with death risk.^3,7,24^ Enrolling patients with both high PISE and high death risks increases clinical trial attrition due to mortality. Thus, a second strategy to boost PPV is to dissociate PISE and death risks, enabling identification of patients likely to survive long enough to develop PISE.

In this study, we aimed to determine if incorporating (i) quantitative PISE biomarkers and (ii) death predictors allow better identification of AIS patients most likely to survive and develop measurable late seizures—ideal candidates for anti-epileptogenic trials.

## Methods

### Study design, setting, and patients

In this *predictive model development* study, we retrospectively collected patient data from Yale-New Haven Hospital by reviewing electronic medical records. We followed the TRIPOD guideline in model development.^25^

We identified a training cohort (admitted from January 1, 2014, to December 31, 2020) for PISE model development and a testing cohort (admitted from January 1, 2021, to July 31, 2022) for internal model validation.

Inclusion criteria were: (1) acute clinical presentation of AIS that correlated with radiologically confirmed infarct, (2) aged ≥18 years, (3) had follow-up notes available >7 days after stroke, (4) had magnetic resonance imaging (MRI) or computed tomography (CT) assessment ≤7 days post-AIS, and (5) underwent continuous EEG (cEEG) monitoring ≤7 days post-AIS. We excluded patients with a history of seizure/epilepsy, brain injuries <5 years pre-AIS (e.g., ischemic stroke, traumatic brain injury, intracerebral/extracerebral hemorrhage), or history of brain diseases known to be highly epileptogenic (e.g., brain tumor).

### Outcomes

PISE was defined as unprovoked clinical/electrographic seizure(s) >7 days after AIS,^26^ which were identified by reviewing clinical notes. Death after AIS was considered a competing event for PISE, which was determined by electronic medical records and obituaries. Analysis endpoints were defined as time from the last known normal datetime to the first >7-day seizure, death, or loss to follow-up.

### Clinical, neuroimaging, and EEG features

We quantified clinical, neuroimaging, and EEG features previously reported to be associated with PISE.^7,16,17,20^

We gathered patient demographic information, past medical history, and in-hospital clinical data, including the five SeLECT variables: admission National Institutes of Health Stroke Scale (NIHSS) score, stroke etiology,^7,27^ early clinical seizures ≤7 days post-AIS, cortical infarct, and middle cerebral artery (MCA) infarct.^7^ Missing NIHSS scores were retrospectively determined by board-certified neurologists (AZ, JAK) based on medical notes.

Infarct volumes were quantified using T2 diffusion-weighted MRI, or non-contrast CT scans if there was no available MRI (appendix p2). Infarct volume was manually traced using Horos (Horos Project, Geneva, Switzerland), and algorithmically computed using RapidAI (iSchemaView, Menlo Park, CA) using ≤620 ×10^-6^ mm^2^/s and manually determined optimal Apparent Diffusion Coefficient (ADC) thresholds.

Automated EEG algorithms were applied to quantify epileptiform abnormality (EA) and background signatures (appendix p3). We defined EA as electrographic seizures (ESZ), lateralized periodic discharges (LPDs), generalized periodic discharges (GPDs), lateralized rhythmic delta activity (LRDA), and epileptiform discharges (EDs).^23^ Using SPaRCNet and Persyst,^28,29^ we classified every consecutive 2-second window of an EEG recording into EA (or subtypes, i.e., ESZ, LPDs, GPDs, LRDA) positive or negative. We then calculated EA (or subtypes) burden defined as the percentage of time containing these abnormal patterns. We selected the 95% highest value as a robust representation of peak 1-hour EA (or subtypes) burden. We also calculated the peak 1-hour ED frequency (i.e., the number of EDs). Using Persyst, we quantified EEG global powers/rhythmicity and power/rhythmicity asymmetries for delta (1-4 Hz), theta (4-8 Hz), alpha (8-13 Hz), beta (13-20 Hz), and total (1-20 Hz) frequency bands. Power/rhythmicity asymmetries were defined as absolute power/rhythmicity differences between left and right hemispheres, divided by global power/rhythmicity, in percentages.

### Statistical analysis

A proportional cause-specific hazards (PH_CS_) model was applied to identify features associated with PISE- or death-specific hazards post-AIS. In the training cohort, we first applied a univariable PH_CS=PISE_ model to identify quantitative clinical, neuroimaging, and EEG features associated with the hazards of PISE (i.e., PISE predictors). PISE predictors with p<0.05 in the bivariable analysis adjusting for the SeLECT score were defined as *PISE biomarkers*. To quantify the additive predictive value of quantitative PISE biomarkers, we trained a random survival forest (RSF)^30^ using SeLECT score and quantitative PISE biomarkers combined and compared its performance against that of the SeLECT. Further, we applied the univariable PH_CS=Death_ model to identify features associated with the death hazard (i.e., death predictor). Univariable proportional subdistribution hazards (PH_SD_) modeling was also performed for a comprehensive interpretation of feature-outcome associations.^31^

With death as a competing event for PISE, a variable that increases PISE-specific hazards may not increase PISE incidence or risk.^31^ Therefore, we applied RSF with competing risk to model the cumulative incidence functions (CIFs) of PISE and death.^32^ The final model, “CRIME_PISE_”, included SeLECT score, quantitative PISE biomarkers, and death predictors to estimate CIFs of both outcomes.

To evaluate model discriminability at x-year post-AIS, we performed time-dependent receiver operating characteristic (ROC) analysis *with* competing events.^33^ To evaluate model calibration at x-year post-AIS, we categorized patients into different risk groups based on their predicted risk scores of outcome(s) at x-year. Then, for each risk group, we compared the average model-predicted CIFs vs the Aalen-Johansen estimates of the CIFs at x-year. We also reported PPV, a clinically relevant metric evaluating the proportion of true PISE patients amongst model-predicted positive cases.^25^ We investigated RSF feature importance at the population level (appendix p13).^32^ We also applied the kernel SHAP (SHapley Additive exPlanations) method to gain quantitative insights into how each feature in RSF contributes to the predicted risk scores for individual patients.^34^

Statistical analyses were performed using R (V4.2.1). R packages, survival, rfsrc, timeROC, and shapper were used for classic survival analysis (e.g., PH_CS_ model, PH_SD_ model, Aalen-Johansen estimation), RSF modeling, time-dependent ROC analysis, and SHAP analysis, respectively.

### Standard Protocol Approvals, Registrations, and Patient Consents

This study was approved by the local Institutional Review Board.

### Data Availability

Anonymized data not published within this article will be made available by request from any qualified investigator.

## Results

Of the 3351 adult AIS patients screened, 280 were eligible and included in the analysis, 53 developed PISE (median [IQR]: 6 [3-13] months from AIS to PISE), 104 died without any seizure (1.2 [0.6-6.7] months from AIS to death), and the remaining were censored (32 [18-47] months follow-up; appendix p4-5). Between the training (n=230) and testing (n=50) cohorts, age (median [IQR], 72 [59-80] vs 72 [60-84] years; p=0.504) and sex (male, 45% vs 58%; p=0.138) were not significantly different (appendix p5). However, the training cohort had lower SeLECT scores (5 [3-5] vs 5 [5-5.8]; p=0.004; appendix p5).

In the training cohort, SeLECT^7^ yielded a one-year area under the ROC curve (AUC_1_) of 0.69 (95% CI, 0.59-0.79), with one-year sensitivity of 90% and specificity of 38% (cutoff: ≥4, appendix p7). Three SeLECT predictors were frequently present in the training cohort: admission NIHSS>4 (76%), cortical involvement (85%), and MCA involvement (84%), resulting in 58% of patients having a SeLECT score of 4 or 5 (Figure 1A). Yet, these patients had widely variable NIHSS at 72 hours, infarct volumes, 1-hour peak EA burdens, and EEG background asymmetries (Figure 1B). Importantly, greater 72h NIHSS (HR_CS=PISE_ [95%CI] Δ3, 1.2 [1.1-1.4], p<0.001), larger infarct (manual Δ10mL, 1.06 [1.04-1.08], p<0.001), greater EA burden (Δ10%, 1.2 [1.1-1.3], p=0.002), greater total power asymmetry (Δ10%, 2.0 [1.4-2.9], p<0.001), and greater rhythmicity asymmetry (Δ10%, 1.3 [1.1-1.6], p=0.01) were significantly associated with increased PISE-specific hazards (Table 1). The significance remains after adjusting for the SeLECT score in bivariable analysis (appendix p9). Indeed, augmenting SeLECT with these quantitative PISE biomarkers significantly improved SeLECT performance amongst patients with ≥4 (but not <4) SeLECT scores (AUC_1_ [95% CI], training out-of-bag, 0.77 [0.66-0.88] vs 0.58 [0.46-0.71]; testing, 0.74 vs 0.63; appendix p12,17).

**Table 1.** Variables associated with increased hazard of epilepsy and/or death after stroke.

|  |  | PISE |  | Death |  |
| --- | --- | --- | --- | --- | --- |
|  |  | HR<sub>CS=PISE</sub><br>(95%CI) | p | HR<sub>CS=Death</sub><br>(95%CI) | p |
| **Demographics and Medical Histories** |  |  |  |  |  |
|  | Age, in 10 years | 0.9 (0.7-1.1) | 0.17 | 1.7 (1.4-2.0) | <0.001 |
|  | Pre-stroke mRS score | 0.9 (0.6-1.3) | 0.54 | 1.4 (1.2-1.7) | <0.001 |
|  | History of Afib, yes | 1.4 (0.8-2.7) | 0.26 | 2.4 (1.6-3.7) | <0.001 |
| **Clinical Assessment** |  |  |  |  |  |
|  | Stroke Etiology (vs small-vessel Occlusion) |  |  |  |  |
|  | Large-artery | 1.5 (0.6-3.6) | 0.34 | 1.0 (0.5-2.0) | 0.94 |
|  | Cardioembolism | 1.1 (0.6-2.1) | 0.69 | 1.9 (1.2-3.0) | 0.01 |
|  | Admission NIHSS score, per 3 units | 1.2 (1.1-1.3) | 0.001 | 1.1 (1.0-1.2) | 0.003 |
|  | 72-hour NIHSS score, per 3 units | 1.2 (1.1-1.4) | <0.001 | 1.2 (1.1-1.3) | <0.001 |
|  | Decompressive Craniectomy, yes | 5.0 (2.4-10.5) | <0.001 | 0.7 (0.3-2) | 0.54 |
| **Stroke Infarct Assessment** |  |  |  |  |  |
|  | Final Infarct Volume, per 10 mL |  |  |  |  |
|  | Manual | 1.1 (1.0-1.1) | <0.001 | 1.0 (1.0-1.0) | 0.05 |
|  | RapidAI, ADC<620 | 1.1 (1.0-1.1) | <0.001 | 1.0 (1.0-1.0) | 0.11 |
|  | RapidAI, Best ADC | 1.1 (1.0-1.1) | <0.001 | 1.0 (1.0-1.0) | 0.09 |
|  | Vessel Territory Involvement, yes |  |  |  |  |
|  | MCA | 4.9 (1.2-20) | 0.03 | 1.3 (0.7-2.4) | 0.38 |
|  | ACA | 2.1 (0.9-4.5) | 0.07 | 2.2 (1.3-3.7) | 0.003 |
|  | Lobar Involvement, yes |  |  |  |  |
|  | Parietal | 2.9 (1.4-6.0) | 0.01 | 1.5 (0.9-2.3) | 0.11 |
|  | Temporal | 2.2 (1.1-4.2) | 0.02 | 1.4 (0.9-2.1) | 0.17 |
| **EEG Assessment** |  |  |  |  |  |
|  | Epileptiform Abnormality |  |  |  |  |
|  | 1-hour Peak EA Burden, in 10 % of 1-hour | 1.2 (1.1-1.3) | 0.002 | 1.0 (0.9-1.1) | 0.91 |
|  | Background Signatures |  |  |  |  |
| | Global Theta Power, in 10 μV | 1.5 (0.3-8.0) | 0.66 | 6.2 (2.3-17) | <0.001 |
| | Global Theta Rhythmicity, in 10 μV | 0.9 (0.6-1.5) | 0.75 | 1.8 (1.4-2.2) | <0.001 |
| | Total Power Asymmetry, Δ10% of global | 2.0 (1.4-2.9) | <0.001 | 1.3 (0.9-1.8) | 0.12 |
| | Total Rhythmicity Asymmetry, Δ10% of global | 1.3 (1.1-1.6) | 0.01 | 1.3 (1.1-1.5) | 0.002 |

HR<sub>CS</sub>=Hazard Ratio for proportional Cause-Specific hazard model. mRS=modified Rankin Scale score. Afib=atrial fibrillation. NIHSS= National Institutes of Health Stroke Scale. ADC=Apparent Diffusion Coefficient. MCA=Middle Cerebral Artery. ACA=Anterior Cerebral Artery. EEG=Electroencephalography. EA=Epileptiform Abnormality.

**Figure 1:**
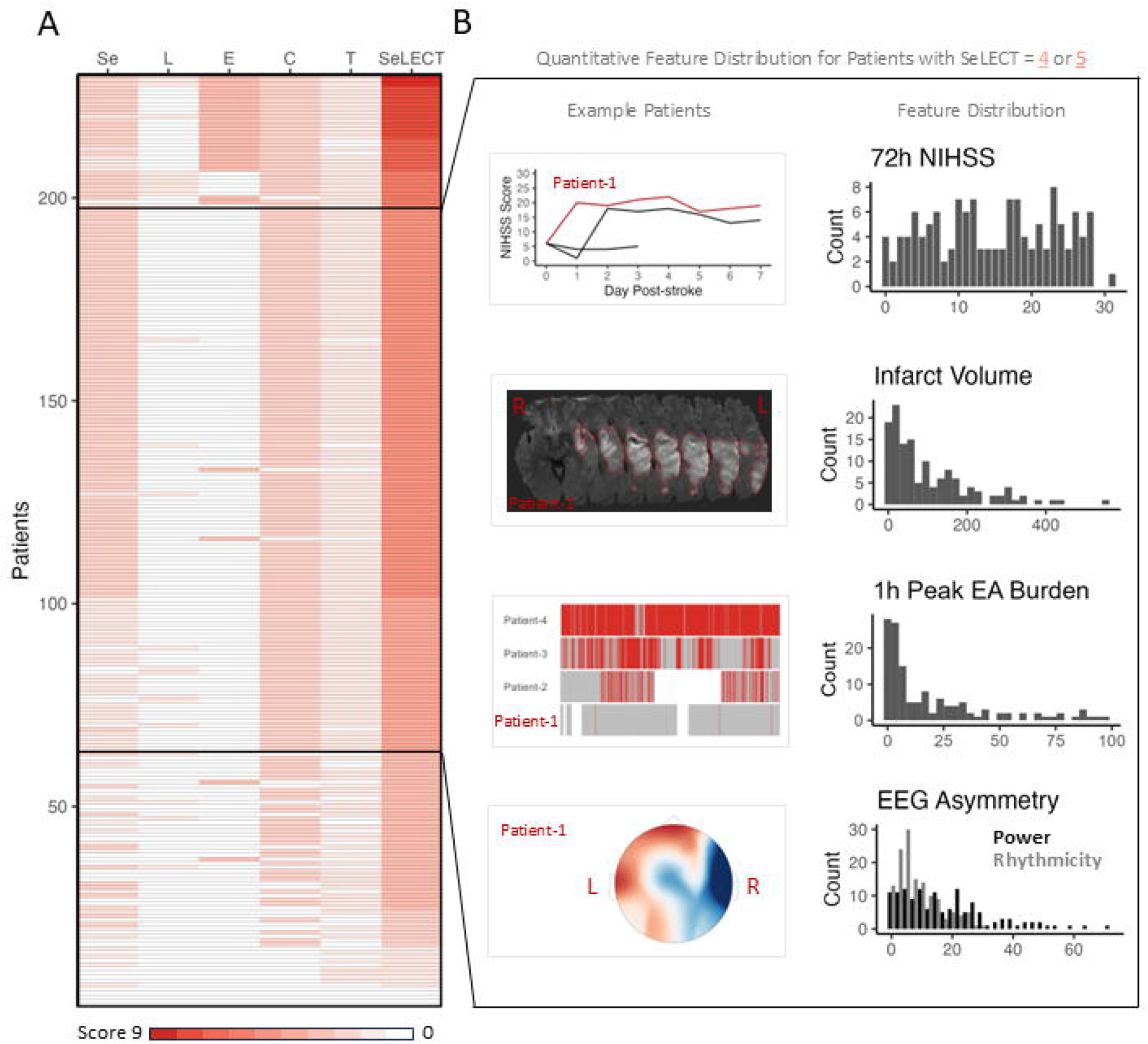
Distributions of SeLECT score and quantitative clinical, neuroimaging, and EEG features for patients in the training cohort. (A) Five SeLECT variables and the total SeLECT score distribution in the training cohort (n=230). Each row represents one patient. Columns from left to right: severity of stroke per admission NIHSS score (4-10, +1; ≥11, +2), large-artery atherosclerosis etiology (yes, +1), early clinical seizure (yes, +3), cortical involvement (yes, +2), territory of MCA (yes, +1), and total SeLECT score. (B) Quantitative clinical, neuroimaging, and EEG feature distributions for patients with SeLECT scores of 4 or 5 (n=134). The left column from top to bottom: example of serial NIHSS score trajectory ≤7 days post-AIS for 3 patients with the same admission NIHSS score; example of manual infarct volume quantification; example of various levels of EA burden during the first 4 hours post-AIS (red – EA, gray – no EA, white – no data); example of EEG total power asymmetry (red – high power, blue – low power). The right column represents histograms of NIHSS scores at 72 hours post-AIS, infarct volume (in mL), 1-hour peak EA burden (in % 1-hour window containing EA), EEG power and rhythmicity asymmetries (in % of global power or rhythmicity). EEG=Electroencephalography. NIHSS=National Institutes of Health Stroke Scale. EA=Epileptiform Abnormality.

Surprisingly, we also found that 31% of patients died at one-year post-AIS, and 84% of these death patients had SeLECT score ≥4 and were falsely predicted to be PISE-positive per SeLECT. Yet, incorporating quantitative PISE biomarkers only minorly corrected these false predictions (40% corrected in training, 35% in testing). The model’s inability to predict whether a patient would experience PISE or death first is evident in Figure 2, where an increased SeLECT score is associated with increased CIFs for not only PISE but death up to a score of 6 (Figure 2).

**Figure 2:**
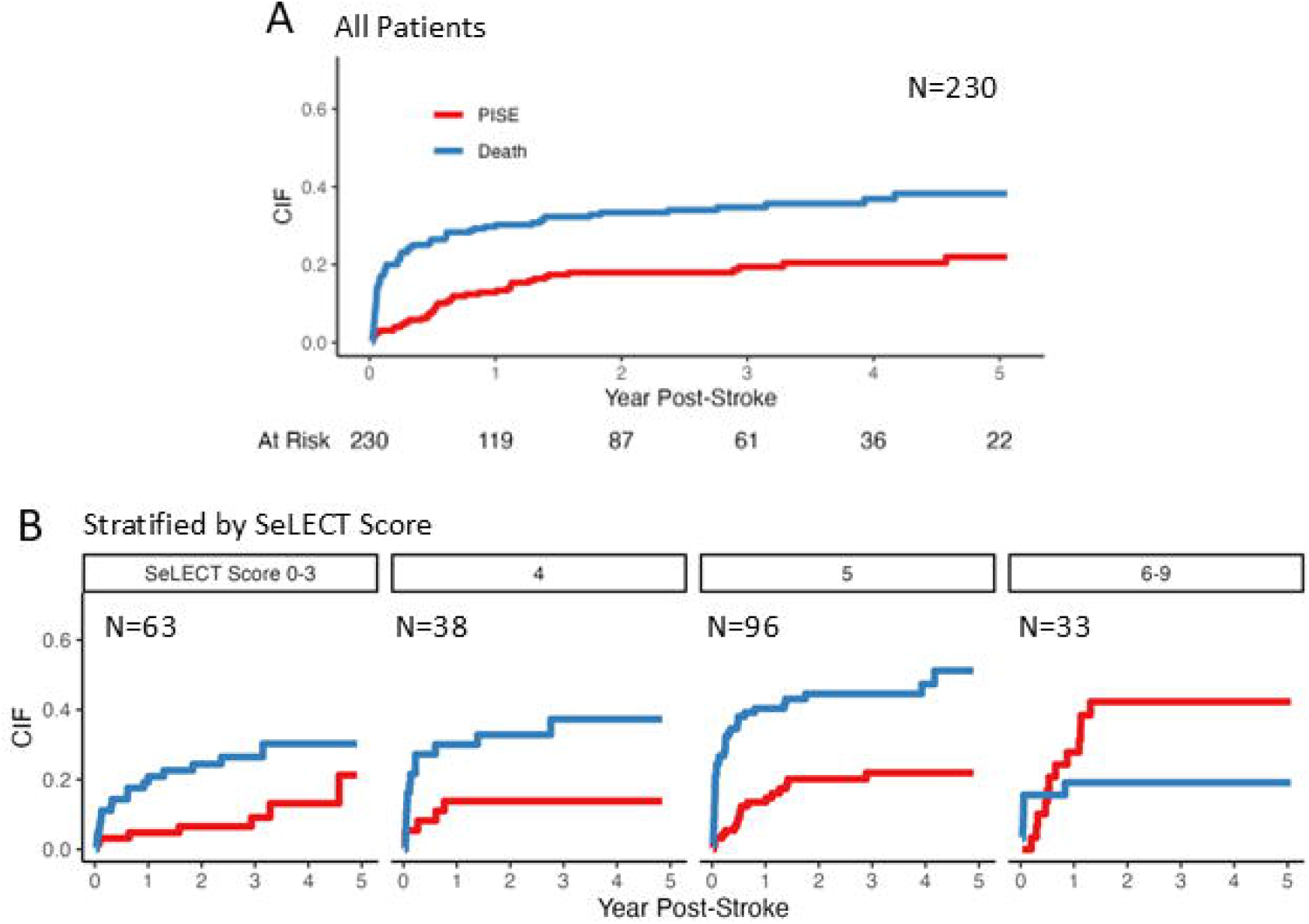
Aalen-Johansen estimates of CIFs of PISE and death for patient subgroups stratified based on the SeLECT score in the training cohort. (A) CIF estimates of PISE (in red) and death (in blue) for all patients in the training cohort (n=230). (B) CIF estimates for patient subgroups with SeLECT scores of 0-3, 4, 5, and 6-9. CIF=Cumulative Incidence Function. PISE=Post-Ischemic Stroke Epilepsy. Death=Death before PISE.

PH_CS=Death_ models show that some PISE predictors in SeLECT and some quantitative PISE biomarkers are also associated with death-specific hazards: admission NIHSS, 72h NIHSS, and rhythmicity asymmetries (Table 1). Additionally, PH_CS=Death_ unveiled death predictors that are associated with only death but not PISE hazards, including older age (HR_CS=Death_ [95%CI] Δ10 years, 1.7 [1.4-2.0], p<0.001), higher pre-stroke modified Rankin Scale score (1.4 [1.2-1.7], p<0.001), atrial fibrillation history (2.4 [1.6-3.7], p<0.001), cardioembolic stroke (1.9 [1.2-3.0], p=0.01), anterior cerebral artery involvement (2.2 [1.3-3.7], p=0.003), global theta power (Δ10 µV, 6.2 [2.3-17], p<0.001), and global total/delta/theta rhythmicity with the largest hazard ratio in the theta band (Δ10 µV, 1.8 [1.4-2.2], p<0.001; Table 1).

Using random survival forest with competing risk framework, we trained the “CRIME_PISE_” (Competing RIsk Mortality & Epilepsy) model which adds quantitative PISE biomarkers and death predictors to the SeLECT score. As compared to SeLECT, CRIME_PISE_ better predicts PISE in both the training (out-of-bag AUC_1_ with competing risk^33^, 0.72 [0.60-0.83] vs 0.65 [0.55-0.75], Figure 3A) and testing (AUC_1_, 0.75 vs 0.57, appendix p14,17) cohorts, but only amongst patients with high (≥4) but not low (<4) SeLECT scores (appendix p14,17). CRIME_PISE_ can also predict death in both training (out-of-bag AUC_1_, 0.79 [0.72-0.85], Figure 3A) and testing (AUC_1_, 0.77) cohorts. CRIME_PISE_ was well-calibrated for both outcomes (appendix p14). Because of the competing risk framework, patients with high predicted risk scores for PISE based on CRIME_PISE_ notably demonstrated increased PISE CIF but decreased death CIF (appendix p14), a trend different from those in SeLECT (Figure 2B).

**Figure 3:**
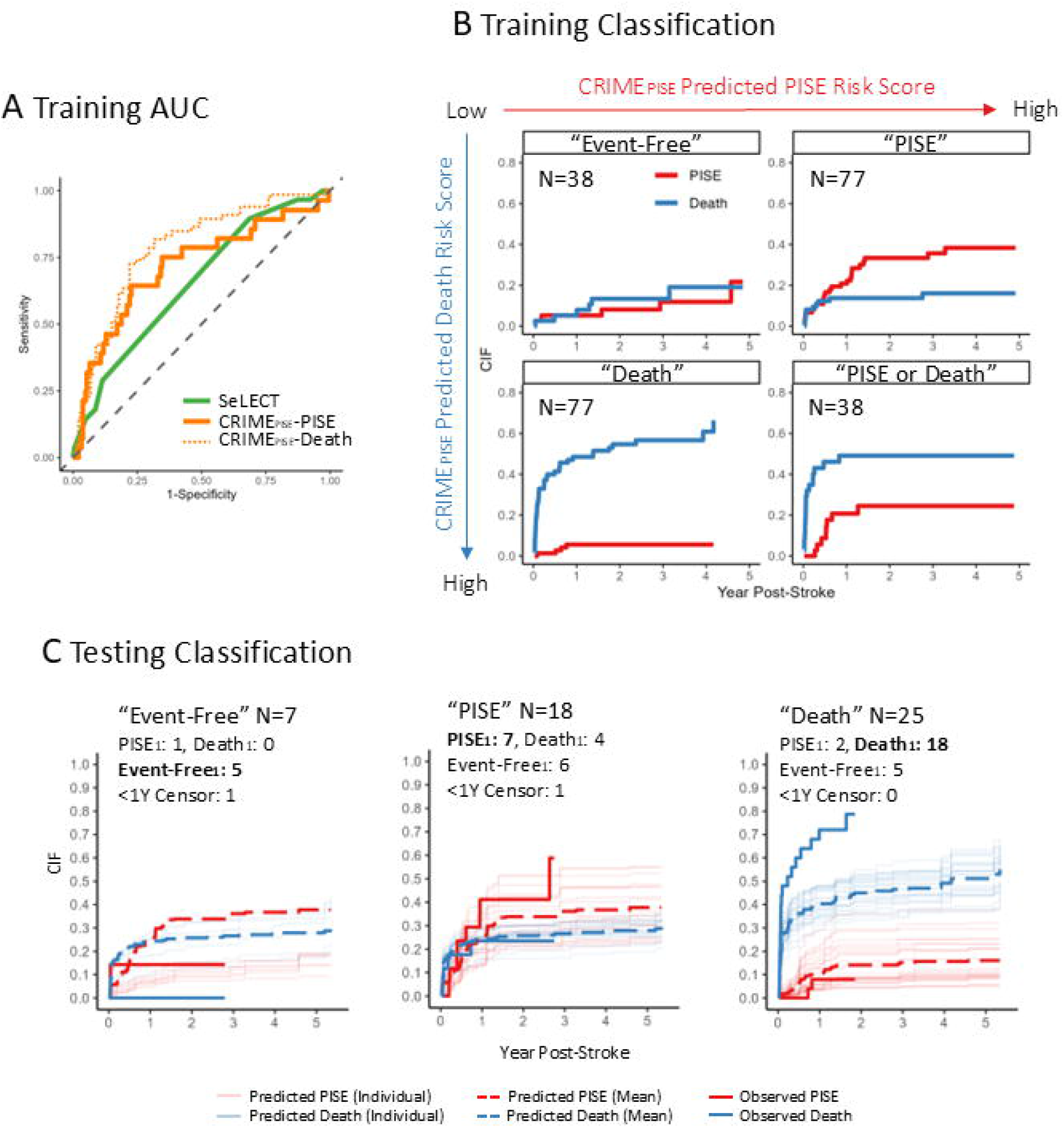
CRIME_PISE_ model performance. (A) First-year out-of-bag ROC (*with* competing risk) curves of the CRIME_PISE_ model (orange) vs the SeLECT model (green) for all patients in the training cohort (n=230). The solid orange line represents the performance for the PISE outcome and the dashed orange line for the death outcome. (B) Same as A but for first-year model calibration analysis. For SeLECT, dots from left to right represent subgroups with SeLECT scores of 0-3 (n=63), 4 (n=38), 5 (n=96), and 6-9 (n=33). For CRIME_PISE_, dots from left to right represent PISE (or death) risk groups corresponding to the quantiles 0-0.27, 0.27-0.44, 0.44-0.86, and 0.86-1, respectively, of the CRIME_PISE_-predicted PISE (or death) risk scores. Error bars represent the 95% confidence interval estimated using bootstrap. The x-axis represents the average of predicted CIFs, and the y-axis represents the Aalen-Johansen estimates of CIFs for each subgroup. For SeLECT, the predicted probabilities of first-year PISE were taken from the SeLECT study. (C) Patients in the training cohort (n=230) were stratified into the 2×2 groups based on whether the predicted out-of-bag PISE (or Death) risk score is above (or below) the population median. “Event-Free”: below-median for both predicted PISE and Death risk scores. “PISE”: above-median for predicted PISE and below-median for predicted Death risk scores. “Death”: above-median for predicted Death and below-median for predicted PISE risk scores. “PISE or Death”: above-median for both predicted PISE and Death risk scores. The red (or blue) curve indicates Aalen-Johansen CIF estimates of PISE (or Death). (D) Patients in the testing cohort (n=50) were stratified using the same threshold as in C. Patients in the “PISE or Death” group were further classified into “PISE” (or “Death”) of the predicted risk score of PISE (or Death) was higher than that of Death (or PISE). Within each CRIME_PISE_ predicted subgroup, the number of of patients who developed first-year PISE (PISE_1_), of patients who died <1-year post-AIS (Death_1_), of patients who had >1-year follow-up indicating seizure-free and death-free (Event-Free_1_), and of patients with <1-year follow-up (<1Y Censor) were reported in the confusion matrix. ROC=Receive Operating Characteristic Curve. PISE=Post-Ischemic Stroke Epilepsy. Death=Death before PISE. CIF=Cumulative Incidence Function.

Utilizing out-of-bag CRIME_PISE_-predicted risk scores for PISE and death, four clinically relevant patient subgroups were defined in the training cohort: “Event-Free” (n=38), “PISE” (n=77), “Death” (n=77), and “PISE or Death” (n=38; Figure 3B). Aalen-Johansen estimates of CIF aligned with the predicted classes (Figure 3B). In the testing cohort, “PISE or Death” patients were further classified as “PISE” if the predicted PISE risk score was greater than that of death, and vice versa. 71% of “Event-Free” patients in the testing cohort were confirmed event-free at one-year post-AIS, 39% of “PISE” patients developed PISE_1_, 72% of “Death” patients had death_1_ (Figure 3C). Compared to SeLECT, PISE had fewer positive predictions (n=18 vs 44), and a larger PPV (39% vs 20%) with a smaller proportion of positive patients who experienced first-year death (22% vs 45%; Figure 3C; Figure 4A Class Label).

**Figure 4:**
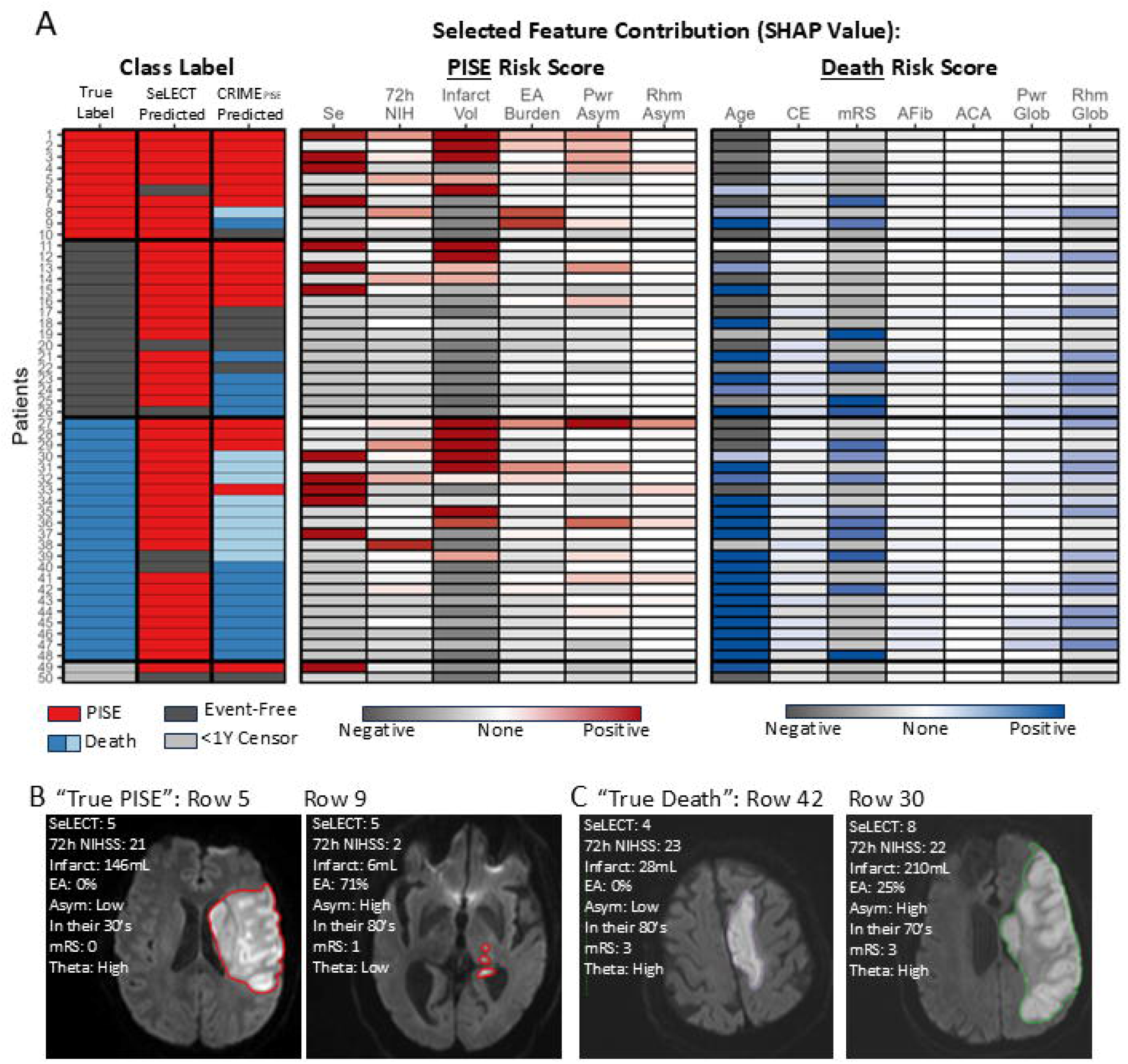
CRIME_PISE_ model classification performance and SHAP-based feature contribution analyses in the testing cohort (n=50) (A) Left panel: Prediction accuracy (first year) of SeLECT and CRIME_PISE_. Each row represents one patient in the testing cohort. Columns from left to right: true label of the patient (outcome evaluated at one-year post-AIS), SeLECT-predicted label using the cutoff of ≥4, CRIME_PISE_-predicted label (light blue indicates patients who had above-median predicted risk scores for both PISE and death, but the risk score of death was higher). Rows were sorted based on the true label and then CRIME_PISE_-predicted PISE risk scores. Middle panel: SHAP value per *PISE-relevant* feature per patient in the CRIME_PISE_ model for the PISE outcome (see appendix p16 for full feature list). Columns from left to right: SeLECT score, NIHSS score at 72 hours post-AIS, infarct volume, 1-hour peak EA burden, EEG total power asymmetry, and EEG total rhythmicity asymmetry. SHAP values were standardized across the entire SHAP-value matrix. More red (or grey) indicates more positive (or negative) contribution of the feature to the CRIME_PISE_-predicted risk scores for PISE; white indicates no contribution. Right panel: same as the middle panel but for death-relevant features for the death outcome in the CRIME_PISE_ model. (B) Examples of PISE_1_ patients with quantitative biomarkers contributed differently to the PISE risk scores. (C) Examples of death_1_ patients. ROC=Receive Operating Characteristic. PISE=Post-Ischemic Stroke Epilepsy. SHAP= Shapley Additive Explanations. NIHSS=National Institutes of Health Stroke Scale. EA=Epileptiform Abnormality. EEG=Electroencephalography. Asym=EEG Power/Rhythmicity Asymmetry. mRS=modified Rankin Scale. Theta=EEG Global Theta Power/Rhythmicity.

SHAP values in the testing cohort provided insights into how quantitative PISE biomarkers and death predictors contribute to the predicted PISE and death risk scores in CRIME_PISE_ at individual patient level (Figure 4A, appendix p16). For PISE patients (rows 1-10, Figure 4A), multimodal (e.g., clinical, neuroimaging, EEG) features contributed *differently* to the predicted PISE risk scores (Figure 4A). For example, one PISE patient had a low, negatively contributing EA burden but a positively contributing 72h NIHSS and infarct volume (Figure 4B, left; another example on the right). For non-PISE patients, CRIME_PISE_ corrected 71% of false predictions made by SeLECT (57% Event-Free patients, rows 11-26; 80% death patients, rows 27-48, Figure 4A). Two examples of SeLECT-positive death patients, one with low, another with high predicted PISE risk score per CRIME_PISE_, had older age, higher pre-AIS mRS score, and high theta rhythmicity positively contributing to their death risks (Figure 4C).

Because CRIME_PISE_ offers added predictive value primarily among patients with SeLECT scores ≥4, we propose a two-stage PISE risk stratification pipeline (Figure 5). In stage one, SeLECT is applied to all AIS patients to identify those at moderate-to-high risk (Figure 5A). These patients with moderate-to-high risk then undergo multimodal neuroimaging and cEEG. In stage two, CRIME_PISE_ is applied to this subgroup to generate individualized predictions of PISE and mortality risk. Based on their risk profile, a patient may fall under a category of “Event-Free”, “PISE” or “Death” (Figure 5B). Their individual patient predicted PISE and mortality risk curves can be plotted and compared to the population based cumulative incidence curves (Figure 5C). Additionally, this framework allows for extraction of personalized feature contributions to highlight distinct contributors to PISE (e.g., large infarcts) and death (e.g., older age) for every individual patient (Figure 5D). The prediction model and example scripts are available in GitHub (https://github.com/jennkim18/CRIME_PISE.git).

**Figure 5.**
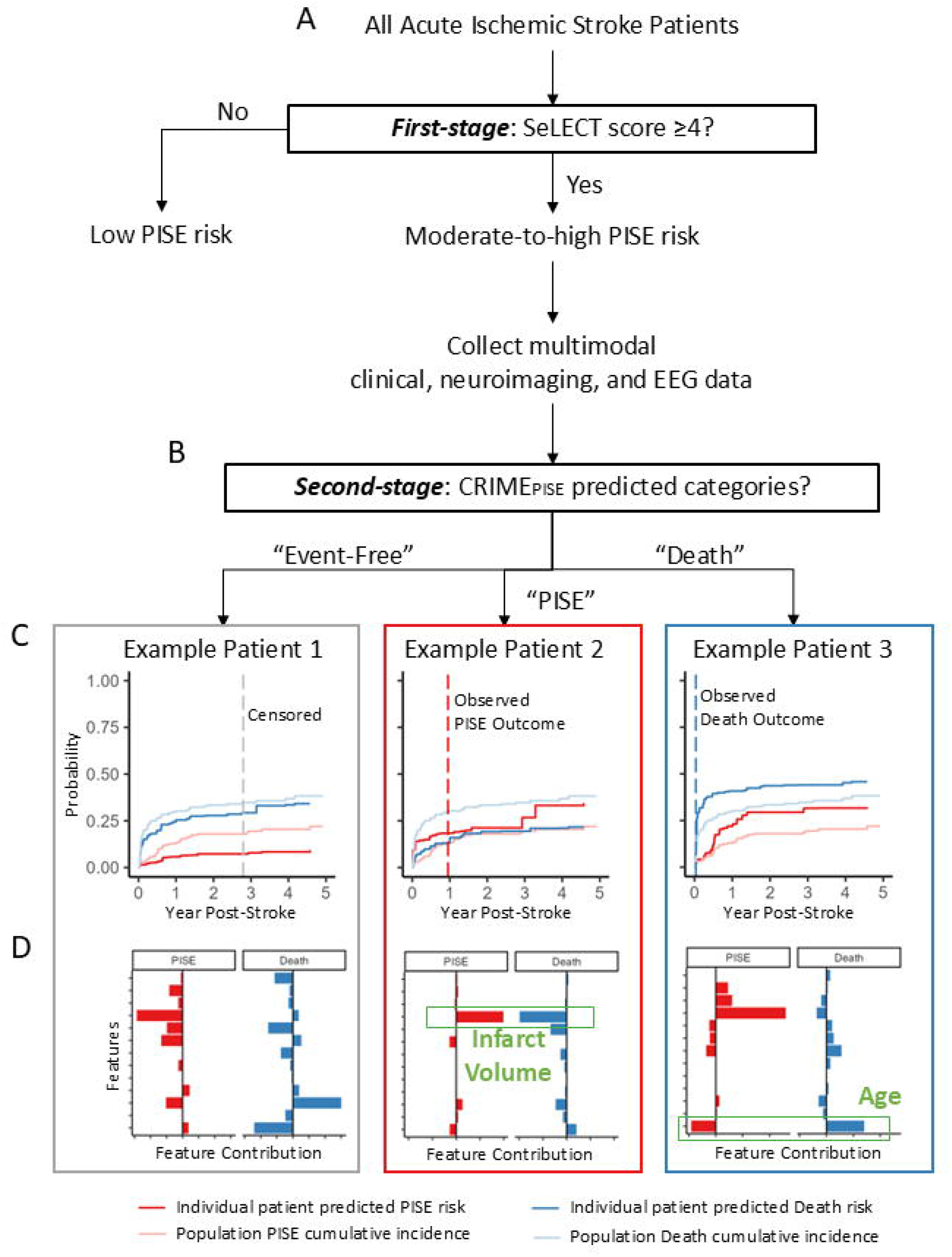
Two-stage risk stratification framework using SeLECT and CRIME_PISE_. This flow diagram illustrates the proposed framework for identifying patients at high risk of developing PISE and surviving long enough for it to manifest. (A) In the first stage, all acute ischemic stroke patients are evaluated using the SeLECT score. Patients with a SeLECT score <4 are predicted to remain PISE-free. (B) Those with a score ≥4 proceed to the second stage, which requires additional clinical resources to obtain multimodal clinical, neuroimaging, and EEG data for CRIME_PISE_-based prediction. CRIME_PISE_ classifies each patient into one of three categories: Event-Free, PISE, or Death. Example patients from each category are shown in gray, red, and blue boxes, respectively. Example Patient 1 was predicted as Event-Free and remained seizure-free up to three years post-stroke. Example Patient 2 was predicted to develop PISE and did so at one year. Example Patient 3 was predicted for Death and died within a month post-stroke. (C) Patient-specific predicted cumulative incidence functions for PISE and death. (D) Contribution of each feature to the predicted risk. For instance, infarct volume contributed most to the PISE risk in Patient 2, while age was the primary contributor to death risk in Patient 3. PISE=Post-Ischemic Stroke Epilepsy. Death=Death before PISE. EEG=Electroencephalography.

## Discussion

CRIME_PISE_ substantially improved upon SeLECT by doubling the PPV for PISE through two strategies.^7–10,35,36^ First, CRIME_PISE_ integrated quantitative PISE biomarkers that provide additional PISE-relevant information beyond the SeLECT score. This enhances differentiation between *non-epileptic* and epileptic AIS patients. Second, CRIME_PISE_ incorporated death predictors to distinguish patients likely to *die without late seizures* from those likely to develop PISE before death. CRIME_PISE_ can be applied conjunctively with SeLECT in antiepileptogenic treatment trials to facilitate targeted enrollments with reduced follow-up costs for non-epileptic patients, and reduced attrition costs due to death (Figure 6A).

**Figure 6:**
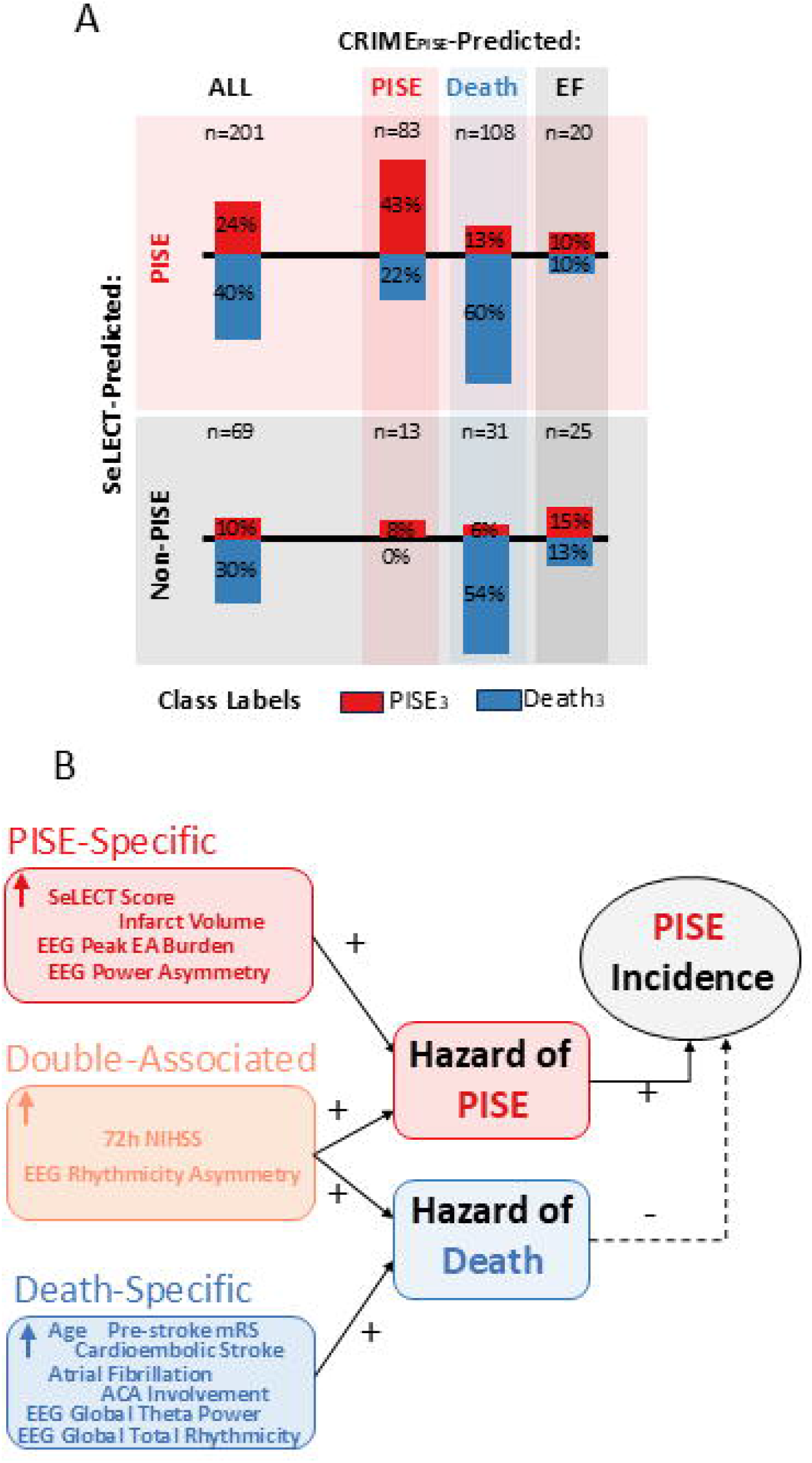
Benefits of CRIME_PISE_. (A) The benefits of using CRIME_PISE_ conjunctively with SeLECT. Two rows represent SeLECT-predicted PISE (n=201) and Non-PISE patients (n=69), respectively. Four columns from left to right represents ALL patients, CRIME_PISE_-predicted PISE, Death, and Event-Free patients, respectively. The length of bar represents the incidence of PISE (red) and Death (blue), in %, which were calculated based on Aalen-Johansen CIF estimates at the third-year post-AIS (PISE3, Death3) within each subgroup. Data were calculated based on the training (out-of-bag) and testing cohort combined. (B) The benefits of considering death in interpreting PISE predictors. PISE-specific (or Death-Specific) predictors represent variables associated with increased hazard for PISE (or Death) but not Death (or PISE), whereas double-associated predictors represent variables associated with increased hazards for both outcomes. The incidence of PISE could be affected by all three classes of predictors. PISE=Post-Ischemic Stroke Epilepsy. Death=Death before PISE. EF=Event-Free. EEG=Electroencephalography. EA=Epileptiform Abnormality. NIHSS=National Institutes of Health Stroke Scale. mRS=modified Rankin Scale score. ACA=Anterior Cerebral Artery.

A notable advantage of CRIME_PISE_ is its incorporation of quantitative PISE biomarkers, including infarct volume, EA burden, EEG background asymmetries, and 72-hour NIHSS score for PISE vs death risk estimation. Infarct size is a widely reported PISE predictor, but remained qualitatively and inconsistently evaluated across studies.^12–15^ Using manual and algorithmic infarct quantification methods, we demonstrated that greater infarct volume was associated with increased hazard and incidence of PISE but not death (i.e., PISE-specific biomarker, Figure 6B). For EEG, we showed that greater EA burden and background power asymmetries were significant PISE-specific biomarkers, even when early clinical seizures did not predict PISE (Figure 6B). This is in accordance with prior studies demonstrating the benefits of regional slowing and EA in PISE prediction. While these studies primarily classified EEG features as binary (present/absent),^16,20^ we incorporated algorithmic, continuous-scale quantification. Hence, our approach can complement these earlier findings and captures gradations of abnormality that may provide additional predictive precision. Together, these findings underscore the promise of post-AIS cEEG monitoring in identifying PISE-relevant subclinical events.^5,16,17,19,20^ Interestingly, EEG rhythmicity asymmetries were associated with increased PISE- and death-specific hazards (i.e., double-associated biomarker), resulting in its non-significant association with the incidence of PISE (Figure 6B). Seventy-two-hour NIHSS was another double-associated biomarker correlated with the incidence of both outcomes. This suggests that stroke severity assessment after acute intervention (e.g., tPA, thrombectomy) provides valuable prognostic information about both PISE and survival,^11,37^ but became an inferior marker if the goal is to distinguish PISE and death. CRIME_PISE_ feature contribution revealed that for PISE patients, the risks for PISE were distributed across these quantitative, multimodal biomarkers. Conversely, for event-free patients, multimodal biomarkers consistently lower the predicted PISE risk, correcting the false positive predictions made by SeLECT for likely non-epileptic AIS patients.

While quantitative neuroimaging and cEEG assessments enhanced PISE prediction, logistical and financial challenges currently limit their application to all AIS patients. We showed that quantitative PISE biomarkers benefited only patients with SeLECT scores ≥4 but not <4 (Figure 6A; appendix p17). This suggests a promise of a two-stage PISE risk stratification framework (Figure 5). Specifically, during the initial risk-stratification phase, SeLECT can be applied to all AIS patients upon admission. Subsequently, only patients exhibiting high SeLECT scores can undergo quantitative neuroimaging and cEEG evaluation that demands more resources (Figure 5). This staged framework offers a cost-effective means of identifying low incident PISE cases among AIS patients.

The second-stage model in the proposed two-stage risk stratification framework will be applied to a selective, more severe AIS patient group with high PISE and mortality rates. Ignoring death as a competing event might result in identifying patients at high risk for PISE when their risk for death is even greater because of the double-associated predictors (Figure 6B). Thus, in prospective anti-epileptogenic treatment trials, if we apply a second-stage model without considering death, the benefits of identifying more PISE patients might be discounted by the attrition costs of enrolling more patients who eventually drop out because of death. By considering the complex interplay of PISE biomarkers and death predictors, CRIME_PISE_ can serve as an appropriate second-stage risk model that distinguishes patients likely to experience PISE before death from those likely to die before developing PISE. This nuanced understanding could support more cost-effective development of anti-epileptogenic treatment trials and contribute to a more accurate prognosis for AIS patients.

Our study has several limitations. Firstly, our cohort is biased toward severe AIS patients who underwent cEEG. To mitigate selection bias, we used the SeLECT score as a baseline assessment and interpreted the benefits of quantitative PISE predictors in relation to the SeLECT model. Yet, we recognize that such bias may affect the generalizability of our model. Secondly, we performed only internal validation. External validation in an independent medical center should be performed using our model. Thirdly, not all patients had imaging and cEEG at similar time points. For patients who did not have an MRI >24 hours post-AIS, their final infarct volume measurement based on <24-hour MRI or >24-hour CT scans may be imperfect. CEEG start-time and monitoring duration varied across patients, leading to an incomplete representation of the true EEG feature value across the entire 7-day post-AIS window. Future prospective studies aligning neuroimaging and cEEG protocols for all patients are needed to address this gap. Fourthly, due to the limited sample size, we focused on modeling the transition probabilities from AIS to PISE or death. Future studies should model the probability of transitioning from PISE to death for a comprehensive understanding of the health state transitions post-AIS. Finally, future studies can refine CRIME_PISE_ with additional neuroimaging, blood, and genetic biomarkers for PISE in the proposed two-stage risk stratification framework.^5^

To conclude, we proposed a PISE prediction model, CRIME_PISE_, which doubles the PPV of SeLECT by augmenting it with quantitative PISE biomarkers and simultaneous death predictions. CRIME_PISE_ significantly outperformed SeLECT only for patients with high but not low SeLECT scores, suggesting the promise of a two-stage risk stratification framework where only patients with high SeLECT scores receive further CRIME_PISE_ evaluations. The proposed two-stage framework would allow anti-epileptogenesis treatment trials to enroll patients who are most likely to experience PISE before death, reducing enrollment, follow-up, and death-associated attrition costs. In the future, when early neuroprotective treatments are available, CRIME_PISE_ might also aid in patient communication and early prognostication of multiple outcomes post-AIS.

## Data Availability

All data produced in the present study are available upon reasonable request to the authors

## Acknowledgements

AFS receives support from the National Institute of Neurological Disorders and Stroke (NINDS, R01NS111022, and R01NS126282). EJG receives support from the NINDS (R01NS117904-01). MBW received funding from the National Institutes of Health (NIH, RF1AG064312, RF1NS120947, R01AG073410, R01HL161253, R01NS126282, R01AG073598, R01NS131347, and R01NS130119), and the National Science Foundation (NSF, 2014431). JAK receives funding from the NINDS (K23NS112596-01A1, R01NS117904, R21NS128641, and R01NS126282), the American Academy of Neurology Clinical Research Training Scholarship, and the Swebilius Foundation.

## Author Contributions

JAK, MBW, KNS, and EJG contributed to the acquisition of funding, acquisition of ethical approval, and protocol development. YC, JAK, and MBW contributed to the study conceptualization and design. YC, ALS, TDS, AZ, LL, AS, and JAK contributed to data collection. YC, JAK, MBW, HS, EJG, JJ, WG, and NP contributed to data processing, statistical analysis, and data interpretation. YC and JAK contributed to writing the manuscript. LJH, HB, SFZ, AFS, KNS, EJG, MBW, and JAK contributed to critical review of the manuscript.

