## Supplement for "Dual-outcome Prediction of Post-Ischemic Stroke Epilepsy and Mortality Using Multimodal Quantitative Biomarkers"

### Stroke infarct volume quantification

For manual infarct volume quantification, we traced the circumference of the infarct for each imaging slice (Figure A1, panel A for MRI, panel C for CT), which would give us the area of the infarct per slice. To obtain the infarct volume, we summed up the cross-sectional areas (i.e., area per slice multiplied by the slice thickness) for all slices. Examples of RapidAI-based infarct volume tracing for the middle slice in panel A using different ADC thresholds are illustrated in panel B. Two automated volumes were computed for each scan – one corresponded to an ADC threshold of ≤620 ×10-6 mm2/s, and the other was based on the optimal ADC threshold (≤620, 680, 700, or 720 10-6 mm2/s) determined by a researcher blind to the manually traced infarct volume.

**Figure A1: Manual and algorithmic infarct volume tracing.**

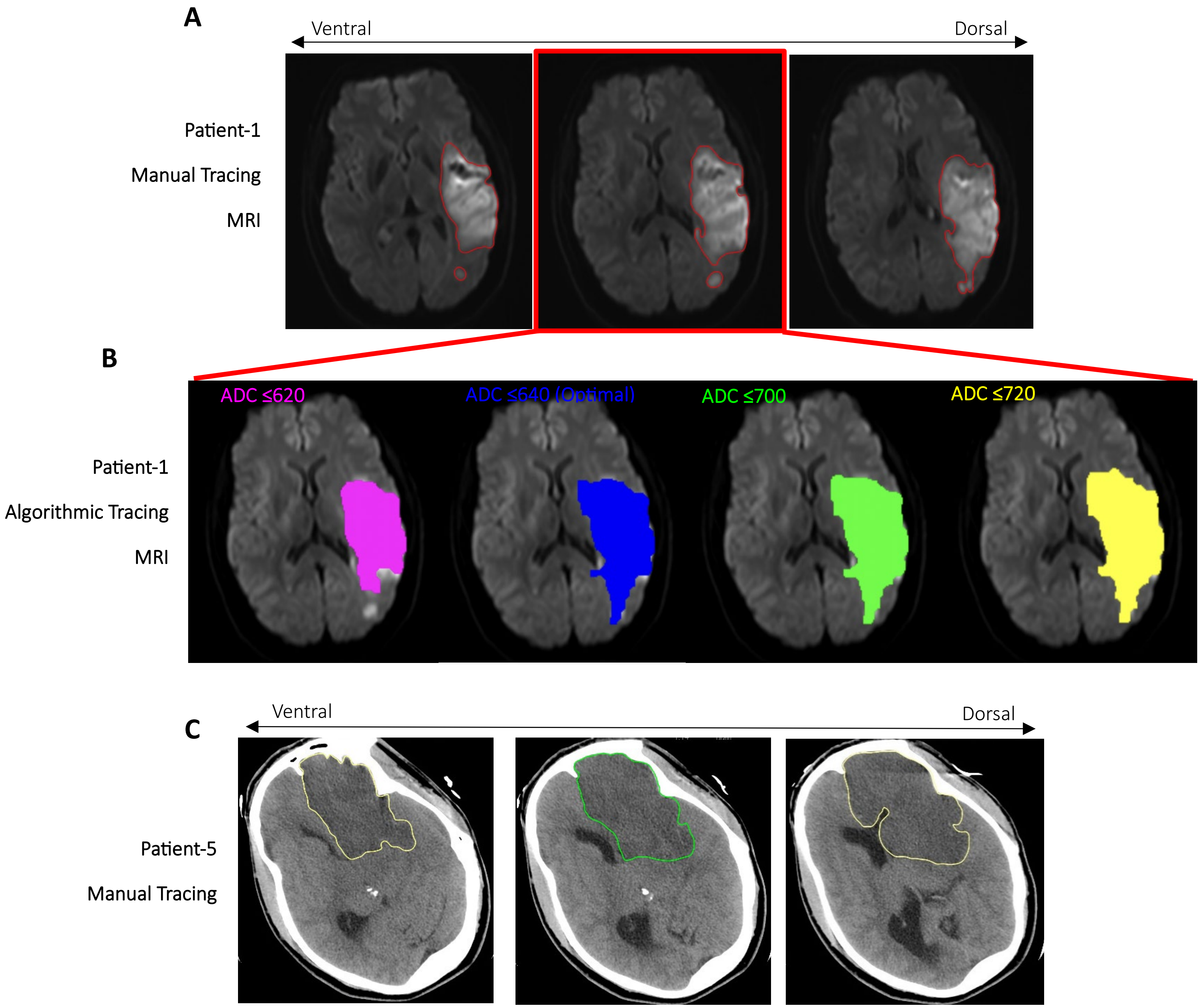

### Electroencephalography (EEG) feature quantification

For each patient, EEG data were recorded using the international 10–20 system. Bipolar montage was used in the following quantitative analysis: Fp1-F7, F7-T3, T3-T5, T5-O1, Fp1-F3, F3-C3, C3-P3, P3-O1, Fp2-F8, F8-T4, T4-T6, T6-O2, Fp2-F4, F4-C4, C4-P4, P4-O2.

(1) 1-hour peak epileptiform abnormality (EA) burden: For each 2-second EEG epoch, SPaRCNet outputs the probabilistic distribution of the target epoch being seizure, LPD, GPD, LRDA, GRDA, or others (e.g., artifacts, normal, etc). The predicted class assigned to each epoch is the one corresponding to the largest probability. Persyst outputs the number of spikes detected for each epoch. A 2-second epoch is defined as EA-positive if it is assigned to a label of seizure, LPD, GPD, LRDA by SPaRCNet, or have >1 spike detected by Persyst. An 1-hour moving average function was applied to the EA feature vectors at 2-second resolution (i.e., 0.5 Hz) to calculate the percentage of time within a 1-hour window that contains EA (i.e., EA burden). The peak burden was defined as the 95% highest EA burden for each patient.

(2) Global power: Persyst outputs background power spectrogram at every 8 seconds. The following parameters were used: time constant = 0.16 seconds, high-frequency filter = 35 Hz, FFT sampling rate = 64 Hz, FFT points per window = 128, FFT window duration = 2 seconds, FFT windows per epoch = 8, overlapped windows = On, FFT smoothing = 3, power range 0-20 Hz. For each patient, global power is defined as the average power of all electrodes across the entire recording.

(3) Global rhythmicity: Persyst outputs background rhythmicity at every 6 seconds. The following parameters were used: time constant = 0.16 seconds, high-frequency filter = 35 Hz, rhythmicity sampling rate = 128 Hz, epoch duration = 3 seconds, epoch step = 2 seconds, frequency range 1-25 Hz. For each patient, global rhythmicity was defined as the average rhythmicity of all electrodes across the entire recording.

(4) Power/rhythmicity asymmetries: for each patient, the left and right hemispheric power/rhythmicity were calculated using the same parameter adopted in calculating the global power/rhythmicity, but were based on averaged left and right electrodes, respectively. The asymmetry was defined as the absolute value of left and right hemispheric power/rhythmicity differences divided by the global power/rhythmicity, and scaled to percentages.

### Patient inclusion and exclusion

**Figure A2: Flow chart of patient inclusion and exclusion for the training (testing) cohorts.**

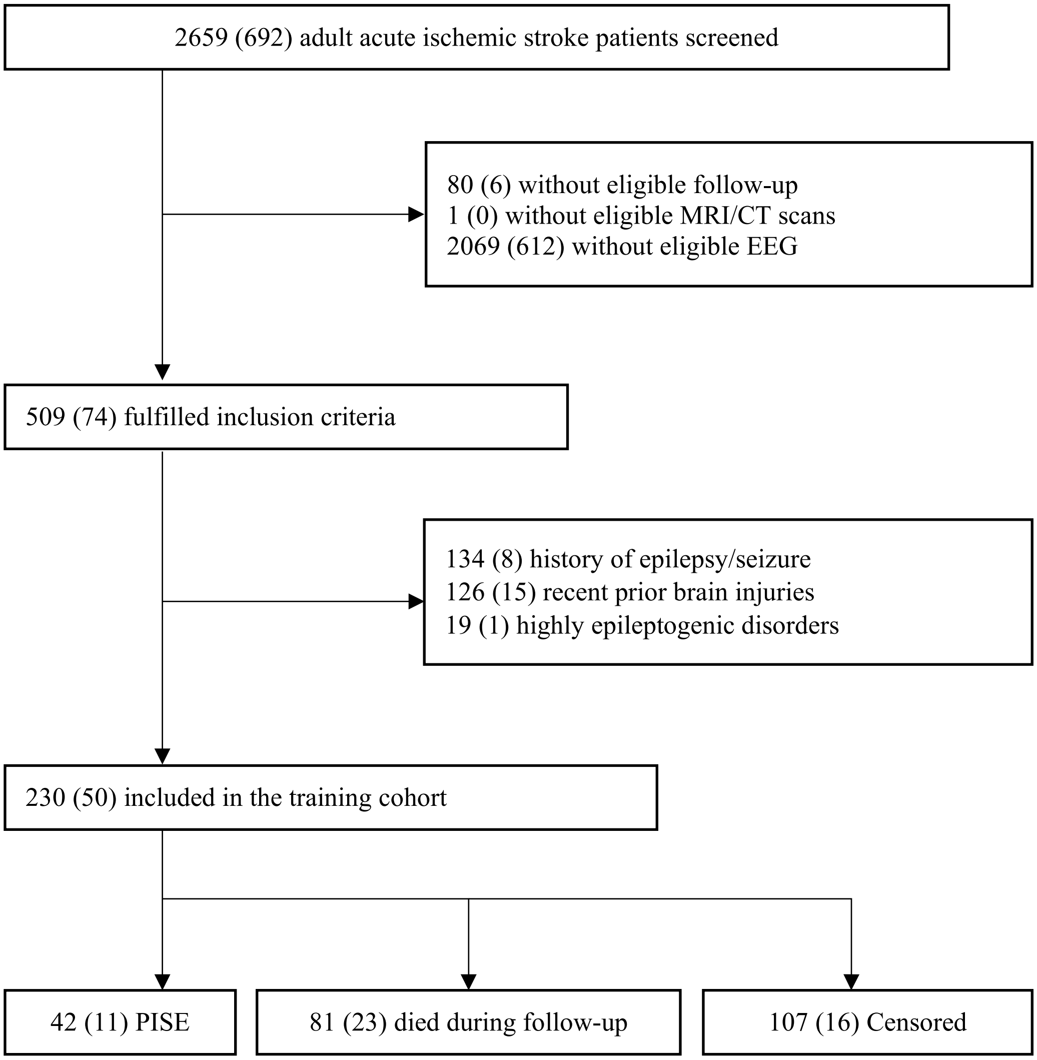

### Patient outcome and baseline characteristics

We applied the same data collection criteria to collect demographics, medical histories, acute stroke interventions, and post-stroke assessment data for the training and the testing patients.

**Table A1: Patient outcome and baseline characteristics**

| | | | Training (n=230) | Testing (n=50) |
|---|---|---|---|---|
| **Chart-review Details** | | | | |
| | Patient admission time range | | Jan 1st 2014 – Dec 31st 2020 | Jan 1st 2021 – July 31st 2022 |
| | Follow-up review cutoff date | | June 2022 | Dec 2023 |
| **Outcomes** | | | | |
| | Months from stroke to the first PISE seizure (PISE) | | 6 (3 – 13) | 6 (3 – 9) |
| | Months from stroke to Death (Death) | | 1.4 (0.6 – 7.3) | 0.9 (0.6 – 3.6) |
| | Month from stroke to the latest follow-up (Censored) | | 36 (18 – 50) | 16 (12 – 23) |
| **SeLECT Score** | | | 5 (3 – 5) | 5 (5 – 5.8) |
| **Demographics and Medical Histories** | | | | |
| | Age, in years | | 72 (59 – 80) | 72 (60 – 84) |
| | Sex | | | |
| | | Male | 104 (45) | 29 (58) |
| | | Female | 126 (55) | 21 (42) |
| | Race | | | |
| | | White or Caucasian | 154 (67) | 31 (62) |
| | | Black or African American | 53 (23) | 8 (16) |
| | | Asian | 5 (2) | 1 (2) |
| | | Pacific Islander | 0 (0) | 2 (4) |
| | | Others or Unknown | 18 (8) | 8 (16) |
| | Ethnicity | | | |
| | | Hispanic or Latino | 19 (8) | 7 (14) |
| | | Non-Hispanic | 209 (91) | 43 (86) |
| | | Unknown | 2 (1) | 0 (0) |
| | Pre-stroke modified Rankin Scale score | | 0 (0 – 1) | 0 (0 – 1) |
| | History of Atrial Fibrillation, yes | | 89 (39) | 20 (40) |
| | History of Remote Brain Injury, yes | | 26 (11) | 8 (16) |
| **Acute Stroke Interventions** | | | | |
| | IV-tPA, yes | | 60 (26) | 15 (30) |
| | Mechanical Thrombectomy, yes | | 68 (30) | 21 (42) |
| | TICI Score | | | |
| | | 0 | 4 (2) | 2 (4) |
| | | 1 | 3 (1) | 0 (0) |
| | | 2A | 8 (4) | 3 (6) |
| | | 2B | 31 (14) | 8 (16) |
| | | 2C | 6 (3) | 6 (12) |
| | | 3 | 16 (7) | 2 (4) |
| | | none | 162 (70) | 29 (58) |
| **Clinical Assessment** | | | | |
| | Hospital Stay, in days | | 12 (7 – 18) | 13 (7 – 19) |
| | Stroke Etiology | | | |
| | | Small-vessel Occlusion | 7 (3) | 2 (4) |
| | | Large-artery Atherosclerosis | 27 (12) | 10 (20) |
| | | Cardioembolism | 112 (49) | 20 (40) |
| | | Others | 26 (11) | 8 (16) |
| | | Undetermined | 58 (25) | 10 (20) |
| | Admission NIHSS score | | 13 (5 – 19) | 17 (12 – 22) |
| | 72-hour NIHSS score | | 11 (4 – 19) | 14 (5 – 22) |
| | Early Clinical Seizures, yes | | 30 (13) | 9 (18) |
| | Hemorrhagic Transformation, yes | | 91 (40) | 32 (64) |
| | Decompressive Craniectomy, yes | | 17 (7) | 5 (10) |
| **Stroke Infarct Assessment** | | | | |
| | Time from Stroke to MRI/CT, in days | | 2 (1 – 3) | 2 (1 – 3) |
| | Final Infarct Volume, in mL | | | |
| | | Manual | 39 (6.0 – 103) | 37 (12 – 150) |
| | | RapidAI, ADC<620 | 23 (0 – 85) | 25 (3 – 136) |
| | | RapidAI, Best ADC | 28 (0 – 95) | 25 (6.0 – 136) |
| | Vessel Territory Involvement, yes | | | |
| | | MCA | 194 (84) | 46 (92) |
| | | ACA | 34 (15) | 10 (20) |

| | | | | |
|---|---|---|---|---|
| | | PCA | 61 (27) | 12 (24) |
| | Infarct Location, yes | | | |
| | | Cortex | 195 (85) | 46 (92) |
| | | White Matter | 210 (91) | 49 (98) |
| | | Basal Ganglia | 132 (57) | 34 (68) |
| | | Cerebellum | 38 (17) | 7 (14) |
| | | Brainstem | 7 (3) | 1 (2) |
| | Lobar Involvement, yes | | | |
| | | Frontal | 167 (73) | 48 (96) |
| | | Parietal | 147 (64) | 37 (74) |
| | | Temporal | 133 (58) | 31 (62) |
| | | Occipital | 92 (40) | 23 (46) |
| **EEG Assessment** | | | | |
| | Time from Stroke to First EEG, in days | | 2 (1 – 3) | 2 (1 – 3) |
| | EEG Monitoring Duration, in hours | | 15 (7 – 24) | 17 (4 – 22) |
| | 1-hour Peak EA Burden, in % of 1-hour | | | |
| | | Total EA | 5 (1 – 19) | 9 (2 – 29) |
| | | ESZ | 0 (0 – 0.2) | 0 (0 – 0.1) |
| | | LPD | 0.2 (0 – 0.9) | 0.2 (0 – 1) |
| | | GPDs | 0 (0 – 0.2) | 0 (0 – 0.2) |
| | | LRDA | 1.4 (0.2 – 7.1) | 0.8 (0.1 – 9.4) |
| | | ED (Frequency) | 41 (7 – 172) | 80 (20 – 406) |
| | 1-hour Peak GRDA Burden | | 1 (0.1 – 5.2) | 0.5 (0 – 5) |
| | Global Power, in μV | | | |
| | | Total (1-20 Hz) | 26 (24 – 30) | 27 (25 – 30) |
| | | Delta (1-4 Hz) | 9 (8 – 10) | 9 (8 – 10) |
| | | Theta (4-8 Hz) | 8 (7 – 9) | 8 (7 – 9) |
| | | Alpha (8-13 Hz) | 7 (6 – 8) | 8 (7 – 9) |
| | | Beta (13-20 Hz) | 7 (6 – 8) | 7 (6 – 8) |
| | Global Rhythmicity, in μV | | | |
| | | Total | 39 (31 – 50) | 44 (34 – 52) |
| | | Delta | 8 (6 – 11) | 8 (6 – 11) |
| | | Theta | 13 (9 – 17) | 14 (12 – 22) |
| | | Alpha | 12 (10 – 16) | 13 (11 – 16) |
| | | Beta | 7 (6 – 9) | 7 (6 – 8) |
| | Power Asymmetry, in % of global power | | | |
| | | Total | 5 (3 – 10) | 8 (5 – 12) |
| | | Delta | 6 (3 – 13) | 8 (3 – 13) |
| | | Theta | 7 (3 – 12) | 9 (5 – 16) |
| | | Alpha | 6 (3 – 10) | 7 (3 – 14) |
| | | Beta | 5 (2 – 8) | 8 (3 – 11) |
| | Rhythmicity Asymmetry, in % of global rhythmicity | | | |
| | | Total | 11 (5 – 22) | 17 (9 – 29) |
| | | Delta | 17 (7 – 30) | 19 (8 – 38) |
| | | Theta | 15 (6 – 27) | 21 (10 – 32) |
| | | Alpha | 9 (4 – 19) | 15 (6 – 24) |
| | | Beta | 8 (4 – 15) | 12 (7 – 20) |

For categorical variables, the number (%) of each category was reported. For numeric variables, the median (interquartile range) was reported. IV-tPA=Intravenous Tissue-type Plasminogen Activator. TICI=Thrombolysis In Cerebral Infarction. NIHSS= National Institutes of Health Stroke Scale. MRI=Magnetic Resonance Imaging. CT= Computed Tomography. ADC=Apparent Diffusion Coefficient. MCA=Middle Cerebral Artery. ACA=Anterior Cerebral Artery. PCA=Posterior Cerebral Artery. EEG=Electroencephalography. EA=Epileptiform Abnormality. ESZ=Electrographic Seizure. LPD=Lateralized Periodic Discharge. GPD=Generalized Periodic Discharge. LRDA=Lateralized Rhythmic Delta Activity. ED=Epileptiform Discharge. GRDA=Generalized Rhythmic Delta Activity.

### SeLECT validation

**Figure A3: Time-dependent AUC without competing risk for the SeLECT model**

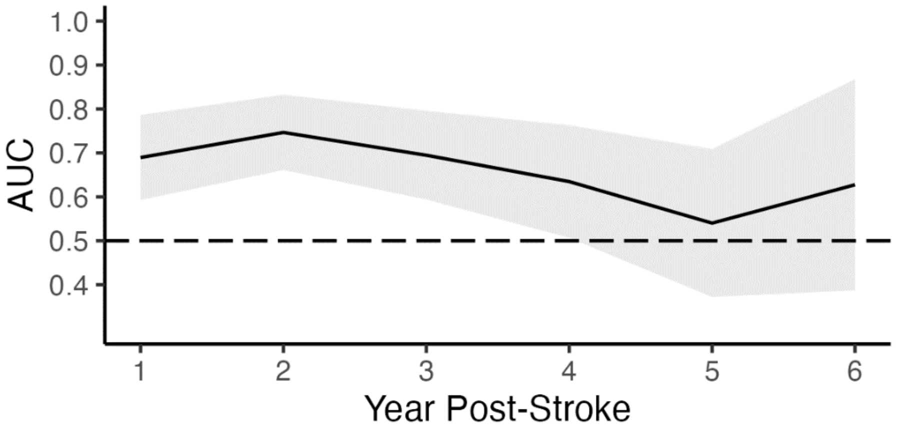

**Table A2: Cutoff analysis without competing risk for the SeLECT model**

| Years | SeLECT Score Cutoff | Sensitivity (%) | Specificity (%) | Positive Predictive Value (%) | Negative Predictive Value (%) |
|---|---|---|---|---|---|
| 1st year post-stroke | 1 | 100 | 4 | 17 | 100 |
| | 2 | 97 | 10 | 18 | 95 |
| | 3 | 97 | 15 | 19 | 97 |
| | 4 | 90 | 38 | 23 | 95 |
| | 5 | 73 | 55 | 25 | 91 |
| | 6 | 30 | 87 | 32 | 86 |
| | 7 | 19 | 91 | 29 | 85 |
| | 8 | 15 | 94 | 34 | 85 |
| | 9 | 4 | 99 | 49 | 84 |
| 2nd year post-stroke | 1 | 100 | 6 | 25 | 100 |
| | 2 | 98 | 13 | 26 | 96 |
| | 3 | 98 | 19 | 28 | 97 |
| | 4 | 90 | 44 | 34 | 93 |
| | 5 | 78 | 64 | 41 | 90 |
| | 6 | 33 | 90 | 50 | 81 |
| | 7 | 22 | 92 | 47 | 79 |
| | 8 | 14 | 93 | 38 | 77 |
| | 9 | 3 | 99 | 43 | 76 |
| 3rd year post-stroke | 1 | 100 | 7 | 28 | 100 |
| | 2 | 98 | 15 | 29 | 96 |
| | 3 | 98 | 17 | 30 | 97 |
| | 4 | 86 | 38 | 34 | 89 |
| | 5 | 76 | 57 | 39 | 87 |
| | 6 | 30 | 88 | 48 | 78 |
| | 7 | 20 | 90 | 42 | 76 |
| | 8 | 12 | 92 | 35 | 74 |
| | 9 | 3 | 100 | 100 | 74 |

### Cox regression analysis

#### Table A4: Estimates of cause-specific hazard and subdistribution hazard for PISE and Death

| | Cox Regression of Cause-Specific Hazard | | | | Cox Regression of Subdistribution Hazard | | | |
|---|---|---|---|---|---|---|---|---|
| | PISE | | Death | | PISE | | Death | |
| | $HR_{CS}$ (95%CI) | p | $HR_{CS}$ (95%CI) | p | $HR_{SD}$ (95%CI) | p | $HR_{SD}$ (95% CI) | p |
| **SeLECT Score** | 1.3 (1.1-1.5) | **0.002** | 1 (0.9-1.2) | 0.48 | 1.3 (1.1-1.5) | **0.002** | 1.0 (0.9-1.1) | 0.66 |
| **Demographics and Medical Histori**es | | | | | | | | |
| Age, in 10 years | 0.9 (0.7-1.1) | 0.17 | 1.7 (1.4-2.0) | **<0.001** | 0.8 (0.7-0.9) | 0.002 | 1.7 (1.5-2.1) | **<0.001** |
| Sex, Male | 1.5 (0.8-2.8) | 0.17 | 0.7 (0.4-1.1) | 0.08 | 1.7 (0.9-3.0) | 0.10 | 0.6 (0.4-1.0) | **0.03** |
| Pre-stroke mRS score | 0.9 (0.6-1.3) | 0.54 | 1.4 (1.2-1.7) | **<0.001** | 0.8 (0.5-1.1) | 0.19 | 1.4 (1.2-1.6) | **<0.001** |
| History of Afib, yes | 1.4 (0.8-2.7) | 0.26 | 2.4 (1.6-3.7) | **<0.001** | 1.0 (0.5-1.9) | 0.98 | 2.2 (1.4-3.4) | **<0.001** |
| History of Remote Brain Injury, yes | 2.0 (0.9-4.4) | 0.11 | 1.7 (0.9-3.1) | 0.11 | 1.7 (0.7-3.9) | 0.22 | 1.4 (0.8-2.5) | 0.26 |
| **Clinical Assessment** | | | | | | | | |
| Stroke Etiology (vs small-vessel Occlusion) | | | | | | | | |
| Large-artery | 1.5 (0.6-3.6) | 0.34 | 1.0 (0.5-2.0) | 0.94 | 1.5 (0.6-3.5) | 0.36 | 0.9 (0.5-1.9) | 0.88 |
| Cardioembolism | 1.1 (0.6-2.1) | 0.69 | 1.9 (1.2-3.0) | **0.01** | 0.9 (0.5-1.6) | 0.69 | 1.9 (1.2-2.9) | **0.01** |
| Others | 1.3 (0.6-3.1) | 0.54 | 0.8 (0.4-1.7) | 0.51 | 1.4 (0.6-3.4) | 0.47 | 0.7 (0.3-1.6) | 0.42 |
| Undetermined | 0.8 (0.4-1.6) | 0.55 | 0.6 (0.3-1.0) | **0.04** | 1.0 (0.5-2.0) | 0.96 | 0.6 (0.3-1.0) | 0.06 |
| Admission NIHSS score, per 3 units | 1.2 (1.1-1.3) | **0.001** | 1.1 (1.0-1.2) | **0.003** | 1.1 (1.0-1.2) | **0.03** | 1.1 (1.0-1.2) | **0.01** |
| 72-hour NIHSS score, per 3 units | 1.2 (1.1-1.4) | **<0.001** | 1.2 (1.1-1.3) | **<0.001** | 1.1 (1.0-1.2) | **0.02** | 1.2 (1.1-1.2) | **<0.001** |
| Early Clinical Seizures, yes | 1.4 (0.6-3.0) | 0.39 | 0.5 (0.2-1.1) | 0.09 | 1.7 (0.8-3.6) | 0.18 | 0.5 (0.2-1.1) | 0.10 |
| Hemorrhagic Transformation, yes | 1.3 (0.7-2.3) | 0.46 | 1.3 (0.8-2) | 0.24 | 1.1 (0.6-2.1) | 0.66 | 1.3 (0.8-2.0) | 0.28 |
| Decompressive Craniectomy, yes | 5.0 (2.4-10.5) | **<0.001** | 0.7 (0.3-2) | 0.54 | 4.5 (2.2-9.3) | **<0.001** | 0.6 (0.2-1.8) | 0.38 |
| **Stroke Infarct Assessment** | | | | | | | | |
| Final Infarct Volume, per 10 mL | | | | | | | | |
| Manual | 1.1 (1.0-1.1) | **<0.001** | 1.0 (1.0-1.0) | 0.05 | 1.0 (1.0-1.0) | **<0.001** | 1.0 (1.0-1.0) | 0.31 |
| RapidAI, ADC<620 | 1.1 (1.0-1.1) | **<0.001** | 1.0 (1.0-1.0) | 0.11 | 1.0 (1.0-1.0) | **<0.001** | 1.0 (1.0-1.0) | 0.37 |
| RapidAI, Best ADC | 1.1 (1.0-1.1) | **<0.001** | 1.0 (1.0-1.0) | 0.09 | 1.1 (1.0-1.1) | **<0.001** | 1.0 (1.0-1.0) | 0.36 |
| Vessel Territory Involvement, yes | | | | | | | | |
| MCA | 4.9 (1.2-20) | **0.03** | 1.3 (0.7-2.4) | 0.38 | 4.3 (1.1-17) | **0.04** | 1.2 (0.7-2.1) | 0.58 |
| ACA | 2.1 (0.9-4.5) | 0.07 | 2.2 (1.3-3.7) | **0.003** | 1.4 (0.6-3.0) | 0.41 | 2.0 (1.1-3.4) | 0.01 |
| PCA | 0.5 (0.2-1.1) | 0.10 | 1.0 (0.6-1.6) | 0.92 | 0.5 (0.2-1.2) | 0.11 | 1.0 (0.7-1.7) | 0.86 |
| Infarct Location, yes | | | | | | | | |
| Cortex | 1.8 (0.7-4.5) | 0.23 | 1.5 (0.8-3.0) | 0.21 | 1.5 (0.6-3.7) | 0.40 | 1.5 (0.8-2.8) | 0.24 |
| White Matter | 1.8 (0.6-5.9) | 0.32 | 2.0 (0.8-4.9) | 0.15 | 1.4 (0.4-4.5) | 0.59 | 1.8 (0.8-4.2) | 0.15 |
| Basal Ganglia | 1.4 (0.7-2.5) | 0.31 | 1.4 (0.9-2.1) | 0.18 | 1.1 (0.6-2.1) | 0.71 | 1.3 (0.9-2.0) | 0.21 |
| Cerebellum | 1.0 (0.4-2.4) | 0.99 | 1.4 (0.8-2.3) | 0.27 | 0.9 (0.4-2.1) | 0.75 | 1.3 (0.8-2.3) | 0.32 |
| Brainstem | 0.6 (0.1-4.5) | 0.63 | 0.3 (0.0-2.4) | 0.28 | 0.8 (0.1-4.6) | 0.76 | 0.4 (0.0-2.8) | 0.34 |
| Lobar Involvement, yes | | | | | | | | |
| Frontal | 1.8 (0.9-3.9) | 0.10 | 1.5 (0.9-2.5) | 0.11 | 1.5 (0.7-3.1) | 0.26 | 1.4 (0.9-2.3) | 0.17 |
| Parietal | 2.9 (1.4-6.0) | **0.01** | 1.5 (0.9-2.3) | 0.11 | 2.3 (1.1-4.8) | **0.02** | 1.3 (0.8-2.1) | 0.23 |
| Temporal | 2.2 (1.1-4.2) | **0.02** | 1.4 (0.9-2.1) | 0.17 | 1.8 (0.9-3.5) | 0.08 | 1.3 (0.8-2.0) | 0.27 |
| Occipital | 1.5 (0.8-2.8) | 0.18 | 0.9 (0.6-1.4) | 0.53 | 1.5 (0.8-2.8) | 0.16 | 0.8 (0.5-1.3) | 0.40 |
| **EEG Assessment** | | | | | | | | |
| 1-hour Peak EA Burden, in 10 % of 1-hour | | | | | | | | |
| Total EA | 1.2 (1.1-1.3) | **0.002** | 1.0 (0.9-1.1) | 0.91 | 1.2 (1.0-1.3) | **0.01** | 1.0 (0.9-1.1) | 0.83 |
| ESZ | 0.5 (0.1-2.6) | 0.43 | 1.2 (0.9-1.7) | 0.27 | 0.5 (0.1-2.1) | 0.38 | 1.2 (0.8-2.0) | 0.35 |
| LPD | 1.2 (1.0-1.5) | **0.02** | 0.9 (0.8-1.2) | 0.62 | 1.2 (1.0-1.4) | **0.04** | 0.9 (0.7-1.2) | 0.53 |
| GPD | 0.4 (0.1-2.1) | 0.29 | 0.6 (0.2-1.6) | 0.31 | 0.5 (0.1-1.7) | 0.24 | 0.7 (0.3-1.3) | 0.23 |
| LRDA | 1.2 (1.1-1.3) | **0.003** | 1.0 (0.9-1.1) | 0.84 | 1.2 (1.1-1.4) | **0.003** | 1.0 (0.9-1.1) | 0.57 |
| ED (Frequency, 100) | 1.1 (1.0-1.2) | **0.002** | 1.1 (1.0-1.1) | **0.01** | 1.1 (1.0-1.1) | 0.06 | 1.1 (1.0-1.1) | 0.05 |
| 1-hour Peak GRDA Burden | 0.9 (0.6-1.1) | 0.30 | 0.8 (0.6-1.0) | 0.08 | 0.9 (0.7-1.2) | 0.49 | 0.8 (0.6-1) | 0.08 |
| Global Power, in 10 µV | | | | | | | | |
| Total (1-20 Hz) | 1.0 (0.5-1.8) | 0.99 | 1.4 (0.9-2.0) | 0.11 | 0.9 (0.5-1.7) | 0.74 | 1.4 (0.9-2.0) | 0.12 |
| Delta (1-4 Hz) | 1.5 (0.3-6.4) | 0.60 | 2.0 (0.8-5.2) | 0.17 | 1.2 (0.2-7.4) | 0.84 | 1.9 (0.7-4.7) | 0.19 |

| | | | | | | | | | | |
|---|---|---|---|---|---|---|---|---|---|---|
| | | Theta (4-8 Hz) | 1.5 (0.3-8.0) | 0.66 | 6.2 (2.3-17) | **<0.001** | 0.7 (0.1-4.3) | 0.66 | 5.7 (1.8-18) | **0.002** |
| | | Alpha (8-13 Hz) | 0.6 (0.1-4.9) | 0.67 | 2.2 (0.6-8.2) | 0.25 | 0.5 (0.1-3.1) | 0.44 | 2.3 (0.6-8.8) | 0.23 |
| | | Beta (13-20 Hz) | 0.4 (0.0-4.8) | 0.49 | 0.7 (0.1-3.7) | 0.67 | 0.5 (0.1-4.8) | 0.58 | 0.8 (0.1-5) | 0.82 |
| | Global Rhythmicity, in 10 µV | | | | | | | | | |
| | | Total | 0.9 (0.8-1.2) | 0.56 | 1.2 (1.0-1.3) | **0.005** | 0.9 (0.8-1.0) | 0.12 | 1.2 (1.1-1.3) | **0.003** |
| | | Delta | 1.2 (0.6-2.2) | 0.64 | 1.7 (1.2-2.5) | **0.006** | 0.9 (0.5-1.6) | 0.74 | 1.7 (1.1-2.6) | **0.02** |
| | | Theta | 0.9 (0.6-1.5) | 0.75 | 1.8 (1.4-2.2) | **<0.001** | 0.7 (0.5-1.1) | 0.09 | 1.8 (1.4-2.3) | **<0.001** |
| | | Alpha | 0.7 (0.4-1.3) | 0.24 | 1.1 (0.8-1.7) | 0.52 | 0.7 (0.4-1.1) | 0.11 | 1.2 (0.8-1.8) | **0.40** |
| | | Beta | 0.4 (0.1-1.6) | 0.20 | 0.9 (0.4-2.1) | 0.74 | 0.5 (0.2-1.4) | 0.18 | 0.9 (0.3-2.8) | 0.92 |
| | Power Asymmetry, Δ10% of global power | | | | | | | | | |
| | | Total | 2.0 (1.4-2.9) | **<0.001** | 1.3 (0.9-1.8) | 0.12 | 1.8 (1.2-2.6) | **0.003** | 1.2 (0.9-1.6) | 0.32 |
| | | Delta | 1.6 (1.2-2.2) | **0.003** | 1.0 (0.7-1.3) | 0.80 | 1.6 (1.2-2.1) | **0.001** | 0.9 (0.7-1.2) | 0.56 |
| | | Theta | 1.7 (1.2-2.4) | **0.002** | 1.3 (1.0-1.7) | 0.06 | 1.5 (1.0-2.1) | **0.03** | 1.2 (0.9-1.5) | 0.17 |
| | | Alpha | 1.9 (1.3-2.7) | **0.001** | 1.4 (1.0-1.8) | **0.02** | 1.5 (1.0-2.3) | **0.03** | 1.2 (0.9-1.6) | 0.12 |
| | | Beta | 1.9 (1.3-2.8) | **0.001** | 1.4 (1.0-1.9) | **0.05** | 1.6 (1.1-2.4) | **0.02** | 1.2 (0.9-1.7) | 0.16 |
| | Rhythmicity Asymmetry, Δ10% of global rhythmicity | | | | | | | | | |
| | | Total | 1.3 (1.1-1.6) | **0.01** | 1.3 (1.1-1.5) | **0.002** | 1.1 (0.9-1.4) | 0.20 | 1.2 (1.0-1.4) | **0.01** |
| | | Delta | 1.2 (1.0-1.4) | **0.01** | 1.1 (1.0-1.3) | **0.02** | 1.1 (1.0-1.3) | 0.16 | 1.1 (1.0-1.2) | **0.06** |
| | | Theta | 1.1 (1.0-1.4) | 0.12 | 1.2 (1.1-1.3) | **0.01** | 1.0 (0.9-1.2) | 0.58 | 1.2 (1.0-1.3) | **0.02** |
| | | Alpha | 1.3 (1.1-1.6) | **0.01** | 1.2 (1.0-1.4) | **0.02** | 1.2 (0.9-1.5) | 0.14 | 1.2 (1.0-1.3) | 0.07 |
| | | Beta | 1.3 (1.1-1.6) | **0.02** | 1.2 (1.0-1.4) | 0.07 | 1.2 (1.0-1.5) | 0.11 | 1.1 (0.9-1.4) | 0.19 |

$HR_{CS}$=Hazard Ratio for proportional Cause-Specific hazard model. $HR_{SD}$=Hazard Ratio for proportional Subdistribution hazard model. mRS=modified Rankin Scale score. Afib=atrial fibrillation. NIHSS= National Institutes of Health Stroke Scale. ADC=Apparent Diffusion Coefficient. MCA=Middle Cerebral Artery. ACA=Anterior Cerebral Artery. PCA=Posterior Cerebral Artery. EEG=Electroencephalography. EA=Epileptiform Abnormality. ESZ=Electrographic Seizure. LPD=Lateralized Periodic Discharge. GPD=Generalized Periodic Discharge. LRDA=Lateralized Rhythmic Delta Activity. ED=Epileptiform Discharge. GRDA=Generalized Rhythmic Delta Activity.

**Table A5 Quantitative biomarkers for PISE derived multivariable proportional cause-specific hazard models**

| | | **Bivariable** | |
|---|---|---|---|
| | | **$aHR_{CS=PISE}$ (95%CI)** | **p-value** |
| **72-hour NIHSS, Δ3 units** | | 1.19 (1.07-1.33) | 0.002 |
| **Infarct Volume, Δ10 mL** | | | |
| | Manual | 1.05 (1.03-1.07) | <0.001 |
| | RapidAI, ADC<620[§] | 1.05 (1.02-1.08) | <0.001 |
| | RapidAI, Best ADC[§] | 1.05 (1.03-1.08) | <0.001 |
| **EEG 1-hour Peak EA Burden, Δ10%** | | | |
| | Total EA | 1.13 (1.02-1.27) | 0.03 |
| | ESZ | 0.41 (0.07-2.30) | 0.31 |
| | LPD | 1.21 (1.01-1.45) | 0.04 |
| | GPD | 0.37 (0.07-1.92) | 0.24 |
| | LRDA | 1.13 (1.00-1.28) | 0.05 |
| | ED (Frequency, Δ100[^]) | 1.09 (1.02-1.16) | 0.009 |
| **EEG Power Asymmetry, Δ10%[*]** | | | |
| | Total (1-20 Hz) | 1.74 (1.20-2.52) | 0.004 |
| | Delta (1-4 Hz) | 1.43 (1.04-1.95) | 0.03 |
| | Theta (4-8 Hz) | 1.55 (1.10-2.19) | 0.01 |
| | Alpha (8-13 Hz) | 1.69 (1.17-2.43) | 0.005 |
| | Beta (13-20 Hz) | 1.66 (1.14-2.42) | 0.009 |
| **EEG Rhythmicity Asymmetry, Δ 10%[*]** | | | |
| | Total | 1.29 (1.04-1.59) | 0.02 |
| | Delta | 1.20 (1.01-1.43) | 0.04 |
| | Theta | 1.13 (0.94-1.35) | 0.21 |
| | Alpha | 1.28 (1.04-1.57) | 0.02 |
| | Beta | 1.25 (1.00-1.56) | 0.05 |

[§]RadpidAI, ADC<620 (or best ADC), RadpidAI computed infarct volume using a threshold of ADC<620 (or the best ADC threshold) for patients with eligible MRI scans (n=200).
[^]The peak number of ED detected within a 1-hour window
[*]10% of global power or rhythmicity

PISE=Post-Ischemic Stroke Epilepsy. HR=Hazard Ratio. aHR=Hazard Ratio adjusted for SeLECT score. CS=Cause-Specific. CI=Confidence Interval. NIHSS= National Institutes of Health Stroke Scale. ADC=Apparent Diffusion Coefficient. EEG=Electroencephalography. EA=Epileptiform Abnormality. ESZ=Electrographic Seizure. LPD=Lateralized Periodic Discharge. GPD=Generalized Periodic Discharge. LRDA=Lateralized Rhythmic Delta Activity. ED=Epileptiform Discharge.

**Table A6: Schoenfeld residuals test of proportional hazard assumption**

| | | | PISE, P-value | Death, P-value |
|---|---|---|---|---|
| **SeLECT Score** | | | 0.980 | 0.160 |
| **Demographics and Medical Histories** | | | | |
| | Age, in years | | 0.248 | 0.232 |
| | Sex, Male | | **0.000** | 0.582 |
| | Pre-stroke modified Rankin Scale score | | 0.310 | 0.350 |
| | History of Atrial Fibrillation, yes | | 0.188 | 0.169 |
| | History of Remote Brain Injury, yes | | 0.191 | 0.064 |
| **Clinical Assessment** | | | | |
| | Stroke Etiology (vs small-vessel Occlusion) | | | |
| | | Large-artery Atherosclerosis | 0.517 | 0.708 |
| | | Cardioembolism | 0.533 | 0.701 |
| | | Others | 0.268 | 0.501 |
| | | Undetermined | 0.216 | 0.572 |
| | Admission NIHSS score | | 0.985 | **0.006** |
| | 72-hour NIHSS score | | 0.259 | **0.005** |
| | Early Clinical Seizures, yes | | 0.646 | 0.345 |
| | Hemorrhagic Transformation, yes | | 0.551 | 0.959 |
| | Decompressive Craniectomy, yes | | 0.521 | 0.201 |
| **Stroke Infarct Assessment** | | | | |
| | Final Infarct Volume, in mL | | | |
| | | Manual | 0.507 | **0.035** |
| | | RapidAI, ADC<620 | 0.064 | 0.098 |
| | | RapidAI, Best ADC | 0.132 | 0.124 |
| | Vessel Territory Involvement, yes | | | |
| | | MCA | 0.501 | 0.464 |
| | | ACA | 0.660 | 0.273 |
| | | PCA | 0.655 | 0.759 |
| | Infarct Location, yes | | | |
| | | Cortex | 0.334 | 0.355 |
| | | White Matter | 0.713 | **0.050** |
| | | Basal Ganglia | 0.648 | **0.004** |
| | | Cerebellum | 0.110 | 0.961 |
| | | Brainstem | 0.153 | 0.459 |
| | Lobar Involvement, yes | | | |
| | | Frontal | 0.115 | 0.080 |
| | | Parietal | 0.436 | 0.474 |
| | | Temporal | 0.571 | 0.309 |
| | | Occipital | 0.591 | 0.909 |
| **EEG Assessment** | | | | |
| | 1-hour Peak EA Burden, in % of 1-hour | | | |
| | | Total EA | 0.779 | 0.296 |
| | | ESZ | 0.693 | 0.156 |
| | | LPD | 0.754 | 0.815 |
| | | GPDs | 0.663 | 0.819 |
| | | LRDA | 0.908 | 0.449 |
| | | ED (Frequency) | 0.196 | 0.725 |
| | 1-hour Peak GRDA Burden | | 0.884 | 0.373 |
| | Global Power, in µV | | | |
| | | Total (1-20 Hz) | 0.142 | 0.873 |
| | | Delta (1-4 Hz) | 0.391 | 0.698 |
| | | Theta (4-8 Hz) | 0.204 | 0.909 |
| | | Alpha (8-13 Hz) | 0.108 | 0.993 |
| | | Beta (13-20 Hz) | 0.078 | 0.871 |
| | Global Rhythmicity, in µV | | | |
| | | Total | 0.287 | 0.533 |

| | | | | |
|---|---|---|---|---|
| | | Delta | 0.971 | 0.168 |
| | | Theta | 0.437 | 0.845 |
| | | Alpha | 0.078 | 0.591 |
| | | Beta | 0.187 | 0.876 |
| | Power Asymmetry, in % of global power | | | |
| | | Total | 0.921 | 0.959 |
| | | Delta | 0.354 | 0.806 |
| | | Theta | 0.969 | 0.998 |
| | | Alpha | 0.537 | 0.706 |
| | | Beta | 0.468 | 0.626 |
| | Rhythmicity Asymmetry, in % of global rhythmicity | | | |
| | | Total | 0.425 | 0.609 |
| | | Delta | 0.905 | 0.244 |
| | | Theta | 0.856 | 0.500 |
| | | Alpha | 0.429 | 0.226 |
| | | Beta | 0.424 | 0.394 |

NIHSS= National Institutes of Health Stroke Scale. ADC=Apparent Diffusion Coefficient. MCA=Middle Cerebral Artery. ACA=Anterior Cerebral Artery. PCA=Posterior Cerebral Artery. EEG=Electroencephalography. EA=Epileptiform Abnormality. ESZ=Electrographic Seizure. LPD=Lateralized Periodic Discharge. GPD=Generalized Periodic Discharge. LRDA=Lateralized Rhythmic Delta Activity. ED=Epileptiform Discharge. GRDA=Generalized Rhythmic Delta Activity.

### The additive benefits of quantitative biomarkers

To determine if selected quantitative biomarkers adds value to the SeLECT socre, we built the "SeQuant" model (SeLECT + Quantitative biomarkers) using random survival forest model which demonstrated several advantages over cox model: (1) modeling non-linear relationships between predictors and outcome, (2) allowing the presence of correlated features that may jointly contribute to the PISE hazard; and (3) estimating model performance using data not included in the training process (i.e., out-of-bag performance).

**Figure A4: SeQuant out-of-bag AUC for 2nd and 3rd years post-stroke**

(A) Distributions of quantitative PISE biomarkers for patients who developed PISE <1-year post-stroke (PISE1, red; n=28) vs patients who had >1-year of follow-up notes indicating seizure-free and death-free (Event-Free1, grey; n=118), stratified by SeLECT scores (all, n=146; 0-3, n=48; 4, n=25; 5, n=50; 6-9, n=23). Top to bottom: NIHSS score at 72 hours post-stroke, infarct volume (mL), 1-hour peak EA burden (%), EEG total power asymmetry (% of total global power), and EEG total rhythmicity asymmetry (% of total global rhythmicity). (B) From top to the bottom: First-, second-, and third-year out-of-bag ROC curve of the SeQuant model (purple) vs the SeLECT model (green) for all patients in the training cohort (n=230). (C) Same as A but only for patients with SeLECT score ≥4 (n=167). ROC=Receiver Operating Characteristic. PISE=Post-Ischemic Stroke Epilepsy. NIHSS=National Institutes of Health Stroke Scale. EA=Epileptiform Abnormality. EEG=Electroencephalography.

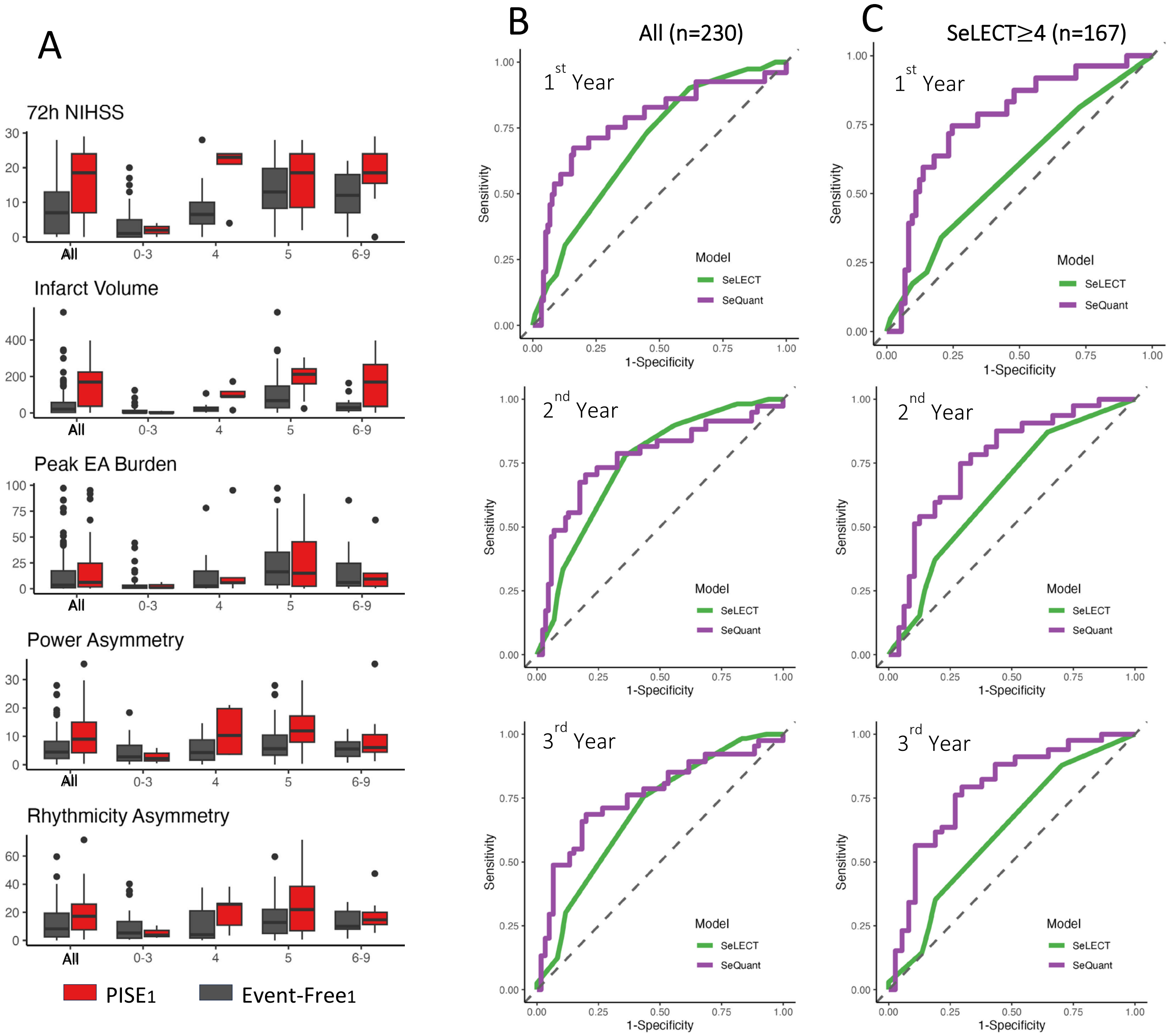

**Table A7: SeQuant cut-off analysis**

| | 1st year post-stroke | | | | 2st year post-stroke | | | | 3rd year post-stroke | | | |
|---|---|---|---|---|---|---|---|---|---|---|---|---|
| **cutoff** | **SEN** | **SPE** | **PPV** | **NPV** | **SEN** | **SPE** | **PPV** | **NPV** | **SEN** | **SPE** | **PPV** | **NPV** |
| **1** | 96 | 8 | 17 | 91 | 97 | 10 | 26 | 92 | 97 | 10 | 28 | 92 |
| **2** | 93 | 31 | 21 | 95 | 88 | 31 | 29 | 89 | 89 | 32 | 32 | 89 |
| **3** | 86 | 47 | 24 | 94 | 84 | 50 | 35 | 91 | 81 | 48 | 36 | 87 |
| **4** | 79 | 60 | 28 | 93 | 79 | 64 | 41 | 90 | 76 | 60 | 41 | 87 |
| **5** | 75 | 64 | 29 | 93 | 73 | 67 | 42 | 89 | **71** | **63** | **41** | **86** |
| **6** | 71 | 70 | 32 | 92 | **70** | **76** | **48** | **89** | 69 | 73 | 48 | 87 |
| **7** | **71** | **77** | **38** | **93** | 67 | 81 | 54 | 89 | 66 | 82 | 56 | 87 |
| **8** | 67 | 81 | 41 | 93 | 56 | 83 | 50 | 85 | 55 | 82 | 52 | 83 |
| **9** | 67 | 84 | 46 | 93 | 56 | 87 | 58 | 86 | 55 | 85 | 57 | 84 |
| **10** | 65 | 85 | 46 | 92 | 54 | 88 | 60 | 86 | 53 | 87 | 59 | 84 |
| **11** | 57 | 86 | 44 | 91 | 49 | 90 | 60 | 85 | 49 | 88 | 60 | 83 |
| **12** | 57 | 88 | 49 | 91 | 49 | 92 | 66 | 85 | 49 | 92 | 68 | 83 |
| **13** | 57 | 88 | 49 | 91 | 49 | 92 | 66 | 85 | 49 | 92 | 68 | 83 |
| **14** | 57 | 89 | 51 | 91 | 49 | 93 | 69 | 85 | 49 | 93 | 73 | 83 |
| **15** | 54 | 90 | 51 | 91 | 46 | 94 | 72 | 85 | 47 | 93 | 72 | 83 |
| **16** | 50 | 92 | 54 | 90 | 43 | 94 | 70 | 84 | 44 | 93 | 70 | 82 |
| **17** | 50 | 92 | 57 | 90 | 41 | 94 | 69 | 83 | 41 | 93 | 69 | 81 |
| **18** | 46 | 93 | 57 | 90 | 38 | 94 | 67 | 83 | 39 | 93 | 68 | 81 |
| **19** | 38 | 94 | 56 | 88 | 30 | 94 | 62 | 81 | 31 | 93 | 63 | 79 |
| **20** | 31 | 95 | 55 | 87 | 25 | 95 | 63 | 80 | 27 | 95 | 66 | 78 |
| **21** | 24 | 95 | 49 | 86 | 20 | 95 | 58 | 79 | 22 | 95 | 62 | 77 |
| **22** | 9 | 97 | 36 | 84 | 10 | 98 | 57 | 77 | 13 | 98 | 74 | 76 |
| **23** | 0 | 97 | 0 | 83 | 3 | 98 | 31 | 76 | 7 | 98 | 61 | 75 |
| **24** | 0 | 97 | 0 | 83 | 3 | 98 | 31 | 76 | 7 | 98 | 61 | 75 |
| **25** | 0 | 98 | 0 | 83 | 0 | 99 | 0 | 76 | 0 | 98 | 0 | 73 |
| **26** | 0 | 98 | 0 | 83 | 0 | 99 | 0 | 76 | 0 | 98 | 0 | 73 |
| **27** | 0 | 99 | 0 | 83 | 0 | 99 | 0 | 76 | 0 | 98 | 0 | 73 |
| **28** | 0 | 99 | 0 | 83 | 0 | 99 | 0 | 76 | 0 | 98 | 0 | 73 |

SEN=Sensitivity. SPE=Specificity. PPV=Positive Predictive Value. NPV=Negative Predictive Value.

**Figure A5: SeQuant feature importance**

Feature importance was defined as the decrease of out-of-bag concordance-index if we randomly assign a daughter node whenever encountering the target feature in the parent node.

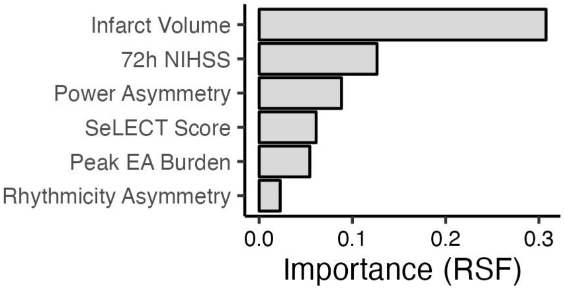

### "CRIMEPISE" model

R package "randomForestSRC" (V3.3.1) was used to build CRIMEPISE, which is a random survival forest with death being a competing event. The outcome include time to one of the three endpoints: PISE, death, or lost-to-followup. The features include age, cardioembolism stroke etiology, pre-stroke modified Rankin Scale score, history of atrial fibrillation, involvement of anterior cerebral artery, global theta power, global theta rhythmicity, SeLECT score, 72h NIHSS score, manually traced infarct volume, peak EA burden, total power asymmetry, and total rhythmicity asymmetry. Modified weighted log-rank splitting rule was applied.

**Figure A6: Second- and third-year out-of-bag ROC and calibration analyses (with competing risk) of the CRIMEPISE model in the training cohort**

(A) Same as Figure 3A-B but for the outcome evaluation at the second- and third-year post-AIS. (B) Same as Figure 3A but only for patients with SeLECT score ≥4 at the second- and third-year post-AIS.

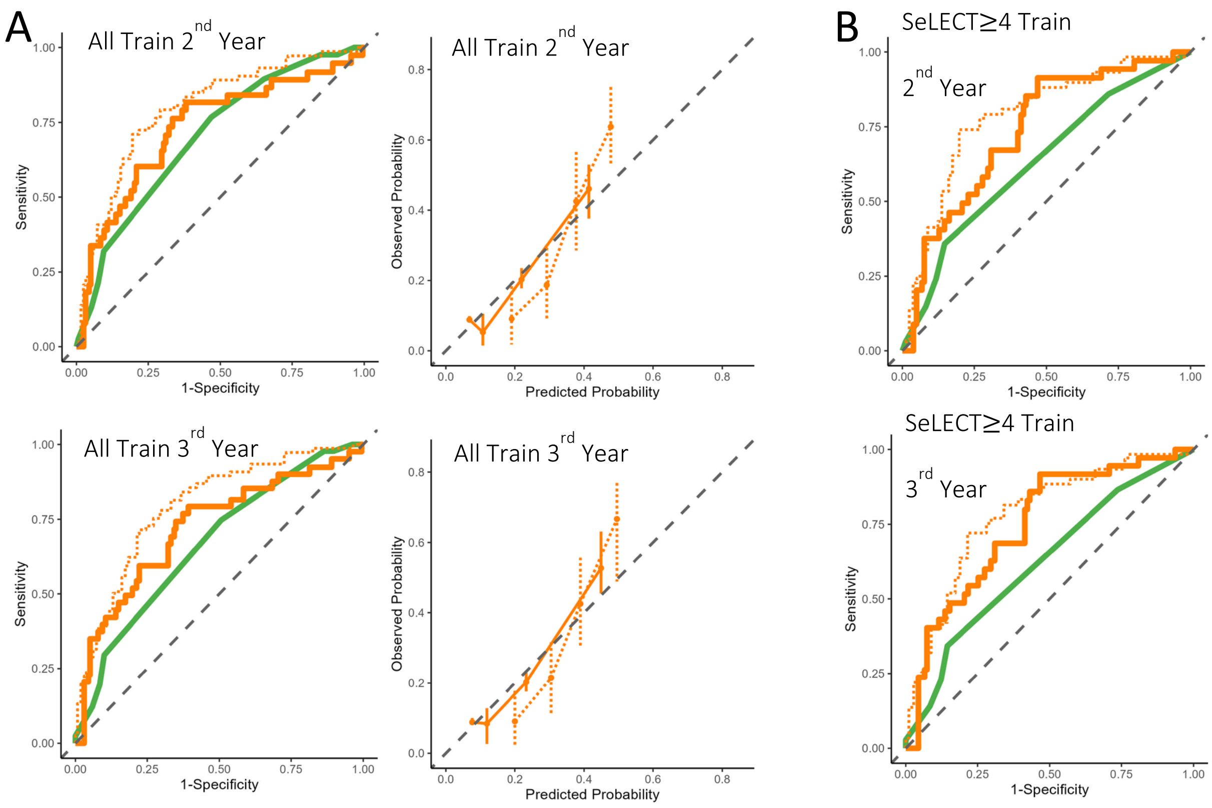

**Figure A7: Aalen-Johansen estimates of CIFs of PISE and death for patient subgroups stratified based on the CRIMEPISE risk score in the training cohort**

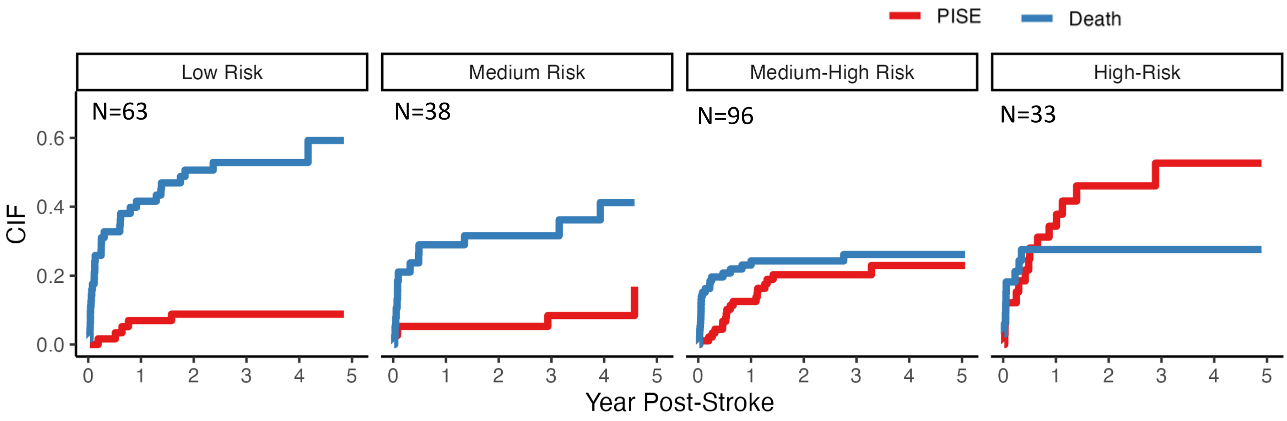

**Table A8: CRIME$_{PISE}$ cut-off analysis**

| | 1st year post-stroke | | | | 2nd year post-stroke | | | | 3rd year post-stroke | | | |
|---|---|---|---|---|---|---|---|---|---|---|---|---|
| **cutoff** | **SEN** | **SPE** | **PPV** | **NPV** | **SEN** | **SPE** | **PPV** | **NPV** | **SEN** | **SPE** | **PPV** | **NPV** |
| **Outcome: PISE** | | | | | | | | | | | | |
| **0.25** | 96 | 1 | 13 | 49 | 97 | 1 | 18 | 49 | 98 | 1 | 19 | 49 |
| **0.37** | 96 | 4 | 13 | 89 | 97 | 4 | 18 | 88 | 98 | 4 | 20 | 88 |
| **0.50** | 93 | 17 | 14 | 94 | 92 | 18 | 20 | 91 | 92 | 17 | 21 | 90 |
| **0.62** | 89 | 29 | 16 | 95 | 89 | 32 | 22 | 93 | 90 | 29 | 24 | 92 |
| **0.75** | 82 | 41 | 17 | 94 | 84 | 44 | 25 | 93 | 82 | 43 | 26 | 91 |
| **0.87** | 79 | 49 | 19 | 94 | 82 | 53 | 28 | 93 | 79 | 53 | 29 | 91 |
| **1.00** | 75 | 58 | 21 | 94 | 79 | 62 | 31 | 93 | 77 | 61 | 32 | 92 |
| **1.12** | 75 | 64 | 23 | 95 | 74 | 68 | 33 | 92 | 72 | 66 | 34 | 91 |
| **1.25** | 64 | 68 | 23 | 93 | 60 | 70 | 31 | 89 | 59 | 68 | 31 | 87 |
| **1.37** | 64 | 72 | 25 | 93 | 60 | 75 | 34 | 90 | 59 | 72 | 34 | 88 |
| **1.50** | 64 | 76 | 28 | 93 | 60 | 78 | 38 | 90 | 59 | 77 | 38 | 89 |
| **1.62** | 57 | 78 | 28 | 93 | 52 | 80 | 37 | 88 | 52 | 79 | 38 | 87 |
| **1.75** | 53 | 81 | 30 | 92 | 49 | 83 | 39 | 88 | 50 | 83 | 41 | 87 |
| **1.87** | 46 | 83 | 29 | 91 | 44 | 86 | 40 | 88 | 45 | 85 | 42 | 86 |
| **2.00** | 46 | 87 | 34 | 92 | 42 | 89 | 45 | 87 | 42 | 89 | 48 | 86 |
| **2.12** | 35 | 90 | 34 | 90 | 34 | 92 | 49 | 86 | 35 | 93 | 54 | 86 |
| **2.25** | 35 | 93 | 43 | 91 | 34 | 95 | 60 | 87 | 35 | 95 | 62 | 86 |
| **2.37** | 35 | 93 | 43 | 91 | 34 | 95 | 60 | 87 | 35 | 95 | 62 | 86 |
| **2.50** | 28 | 95 | 44 | 90 | 23 | 95 | 51 | 85 | 25 | 95 | 54 | 84 |
| **2.62** | 21 | 96 | 45 | 89 | 18 | 97 | 56 | 84 | 21 | 97 | 62 | 83 |
| **2.75** | 7 | 97 | 24 | 88 | 8 | 98 | 41 | 83 | 7 | 97 | 37 | 81 |
| **2.87** | 4 | 97 | 14 | 87 | 5 | 98 | 32 | 82 | 5 | 97 | 29 | 81 |
| **3.00** | 4 | 98 | 20 | 87 | 3 | 98 | 18 | 82 | 2 | 97 | 16 | 80 |
| **3.12** | 0 | 99 | 0 | 87 | 0 | 99 | 0 | 82 | 0 | 98 | 0 | 80 |
| **3.25** | 0 | 99 | 0 | 87 | 0 | 99 | 0 | 82 | 0 | 98 | 0 | 80 |
| **Outcome: Death** | | | | | | | | | | | | |
| **0.25** | 96 | 1 | 13 | 49 | 97 | 1 | 18 | 49 | 98 | 1 | 19 | 49 |
| **0.37** | 96 | 4 | 13 | 89 | 97 | 4 | 18 | 88 | 98 | 4 | 20 | 88 |
| **0.50** | 93 | 17 | 14 | 94 | 92 | 18 | 20 | 91 | 92 | 17 | 21 | 90 |
| **1.00** | 98 | 4 | 30 | 87 | 99 | 5 | 34 | 88 | 99 | 7 | 36 | 91 |
| **1.14** | 98 | 10 | 32 | 94 | 99 | 13 | 36 | 95 | 99 | 14 | 38 | 95 |
| **1.29** | 98 | 15 | 33 | 96 | 97 | 17 | 37 | 93 | 97 | 19 | 39 | 93 |
| **1.43** | 98 | 23 | 35 | 97 | 97 | 26 | 40 | 95 | 97 | 26 | 41 | 95 |
| **1.57** | 94 | 28 | 36 | 92 | 93 | 30 | 40 | 90 | 93 | 31 | 42 | 90 |
| **1.71** | 91 | 37 | 38 | 91 | 90 | 39 | 43 | 89 | 91 | 42 | 45 | 90 |
| **1.86** | 89 | 45 | 41 | 91 | 89 | 47 | 46 | 90 | 90 | 50 | 49 | 90 |
| **2.00** | 89 | 50 | 43 | 92 | 89 | 52 | 48 | 90 | 90 | 53 | 50 | 90 |
| **2.14** | 85 | 58 | 46 | 90 | 83 | 59 | 51 | 88 | 84 | 60 | 53 | 88 |
| **2.29** | 82 | 67 | 51 | 90 | 79 | 69 | 56 | 87 | 78 | 70 | 58 | 86 |
| **2.43** | 74 | 74 | 55 | 87 | 72 | 76 | 61 | 85 | 71 | 77 | 62 | 83 |
| **2.57** | 63 | 81 | 58 | 84 | 63 | 84 | 66 | 82 | 62 | 82 | 64 | 80 |
| **2.71** | 54 | 83 | 57 | 81 | 53 | 86 | 65 | 78 | 53 | 84 | 64 | 77 |
| **2.86** | 42 | 90 | 63 | 79 | 41 | 91 | 70 | 75 | 39 | 90 | 68 | 74 |
| **3.00** | 35 | 92 | 64 | 77 | 34 | 93 | 72 | 74 | 33 | 93 | 71 | 72 |
| **3.14** | 23 | 94 | 64 | 74 | 23 | 97 | 78 | 72 | 22 | 97 | 80 | 70 |
| **3.29** | 17 | 97 | 72 | 73 | 16 | 98 | 77 | 70 | 16 | 98 | 82 | 69 |
| **3.43** | 12 | 98 | 71 | 72 | 12 | 98 | 80 | 69 | 12 | 99 | 90 | 68 |
| **3.57** | 3 | 99 | 49 | 71 | 3 | 98 | 47 | 67 | 3 | 99 | 66 | 66 |
| **3.71** | 1 | 99 | 48 | 70 | 1 | 99 | 44 | 67 | 1 | 100 | 100 | 66 |

SEN=Sensitivity. SPE=Specificity. PPV=Positive Predictive Value. NPV=Negative Predictive Value.

**Figure A8: CRIME$_{PISE}$ feature contribution**

CRIME$_{PISE}$ feature contribution for all features and for both outcomes (full version of Figure 5A). Left panel: Prediction accuracy (first year) of SeLECT and CRIME$_{PISE}$. Each row represents one patient in the testing cohort. Red denotes PISE, dark blue denotes Death, light blue denotes PISE or Death but the CIF of Death > CIF of PISE and were thus classified as Death, dark grey denotes event-free, and light-grey denotes <1 year of follow-up. Rows were sorted based on the true label and then CRIME$_{PISE}$-predicted risk scores for PISE. Middle panel: SHAP value (outcome=PISE) per feature per patient in the CRIME$_{PISE}$ model. More red (or grey) indicates more positive (or negative) contribution of the feature to the CRIME$_{PISE}$-predicted risk scores for PISE; white indicates no contribution. Columns from left to right: age, cardioembolism etiology, modified Rankin Scale score, atrial fibrillation, anterior cerebral artery involvement, global theta power, global theta rhythmicity, SeLECT score, NIHSS score at 72 hours post-stroke, infarct volume, 1-hour peak EA burden, EEG total power asymmetry, and EEG total rhythmicity asymmetry. SHAP values were standardized across the entire SHAP-value matrix. Right panel: same as middle but for the outcome of Death. More blue (or grey) indicates more positive (or negative) contribution of the feature to the CRIME$_{PISE}$-predicted risk scores for Death; white indicates no contribution.

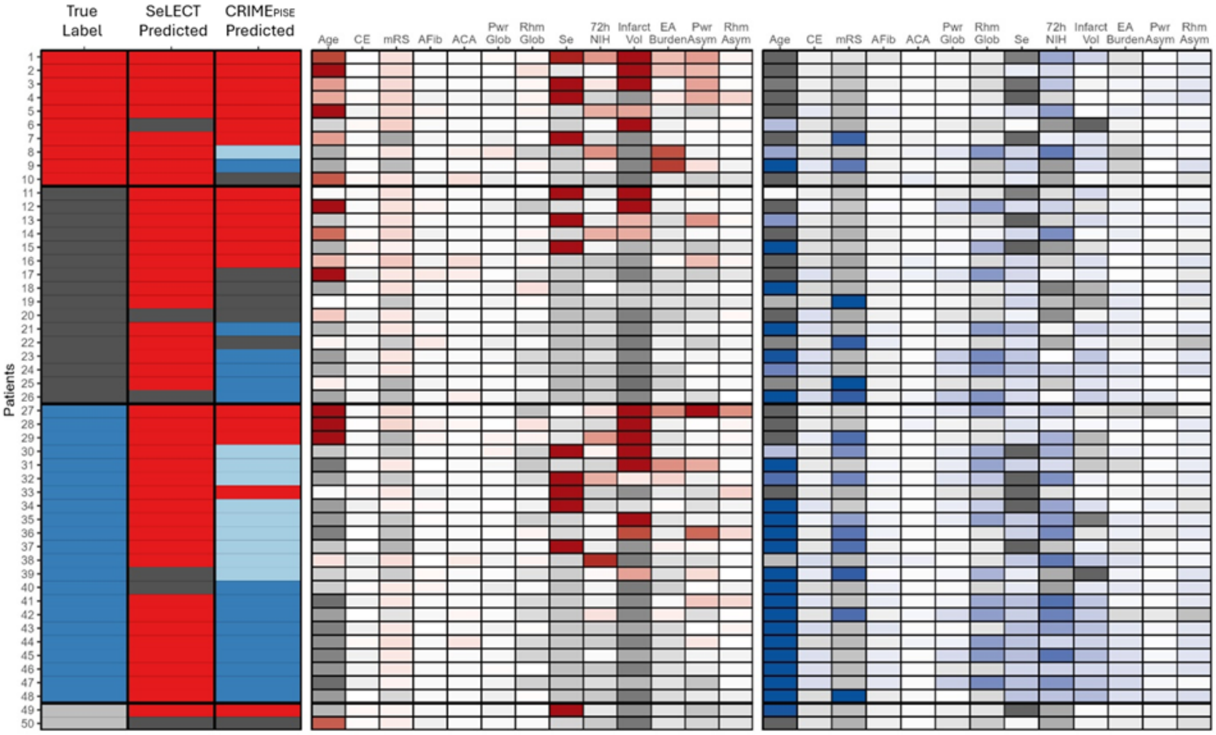

### SeLECT, SeQuant, and CRIME$_{PISE}$ Performance Comparison

**Table A9: Time-dependent AUC comparison**

| | **Without Competing Risk** | | | **With Competing Risk** | | | |
|---|---|---|---|---|---|---|---|
| | **SeLECT** | **SeQuant** | | **SeLECT** | **CRIME$_{PISE}$ PISE** | | **CRIME$_{PISE}$ Death** |
| **Dataset** | AUC (95% CI) | AUC (95% CI) | p | AUC (95% CI) | AUC (95% CI) | p | AUC (95% CI) |
| **Training, All (n=230)** | | | | | | | |
| **1st year** | .69 (.59 – .79) | .77 (.66 – .88) | 0.06 | .65 (.55 – .75) | .72 (.60 – .83) | 0.13 | .79 (.72 – .85) |
| **2nd year** | .75 (.66 – .83) | .77 (.67 – .87) | 0.33 | .70 (.62 – .79) | .73 (.63 – .83) | 0.26 | .80 (.73 – .86) |
| **3rd year** | .69 (.59 – .80) | .76 (.65 – .86) | 0.11 | .67 (.58 – .76) | .72 (.62 – .82) | 0.12 | .80 (.73 – .86) |
| **Training, SeLECT Score≥4 (n=167)** | | | | | | | |
| **1st year** | .58 (.46 – .71) | .77 (.66 – .88) | **0.004** | .58 (.46 – .70) | .72 (.61 – .83) | **0.02** | .76 (.68 – .84) |
| **2nd year** | .64 (.53 – .75) | .76 (.66 – .87) | **0.03** | .63 (.53 – .73) | .74 (.64 – .83) | **0.04** | .79 (.71 – .87) |
| **3rd year** | .62 (.50 – .74) | .78 (.69 – .89) | **0.008** | .62 (.53 – .72) | .75 (.65 – .84) | **0.02** | .78 (.70 – .86) |
| **Training, SeLECT Score<4 (n=63)** | | | | | | | |
| **1st year** | .55 (.27 - .82) | .36 (.01 - .70) | n/a | .52 (.23 - .82) | .33 (.03 - .63) | n/a | .87 (.78 - .97) |
| **2nd year** | .61 (.40 - .81) | .26 (.00 - .53) | n/a | .59 (.35 - .82) | .26 (.03 - .50) | n/a | .81 (.69 - .94) |
| **3rd year** | .66 (.50 - .81) | .32 (.07 - .58) | n/a | .63 (.46 - .81) | .32 (.11 - .54) | n/a | .83 (.71 - .95) |
| **Testing, All (n=50)** | | | | | | | |
| **1st year** | .60 | .80 | n/a | .57 | .75 | n/a | .76 |
| **2nd year** | .57 | .73 | n/a | .57 | .73 | n/a | .85 |
| **Testing, SeLECT Score≥4 (n=44)** | | | | | | | |
| **1st year** | .63 | .64 | n/a | .59 | .72 | n/a | .78 |
| **2nd year** | .78 | .74 | n/a | .60 | .71 | n/a | .83 |
| **Testing, SeLECT Score<4 (n=6), not evaluated due to small sample** | | | | | | | |

AUC = Area Under the receiver operating characteristic Curve
